# Genomic epidemiology and longitudinal sampling of ward wastewater environments and patients reveals complexity of the transmission dynamics of bla_KPC_-carbapenemase-producing Enterobacterales in a hospital setting

**DOI:** 10.1101/2021.11.26.21266267

**Authors:** N Stoesser, R George, Z Aiken, HTT Phan, S Lipworth, DH Wyllie, TP Quan, AJ Mathers, N De Maio, AC Seale, DW Eyre, A Vaughan, J Swann, TEA Peto, DW Crook, J Cawthorne, A Dodgson, AS Walker, TRACE Investigators’ Group

**Affiliations:** Nuffield Department of Medicine, University of Oxford, Oxford, United Kingdom; NIHR Health Protection Research Unit in Healthcare Associated Infections and Antimicrobial Resistance at University of Oxford in partnership with Public Health England, Oxford, United Kingdom; Manchester University NHS Foundation Trust, Manchester, United Kingdom; University of Southampton, Southampton, United Kingdom; United Kingdom Health Security Agency, United Kingdom; University of Virginia, Charlottesville, United States of America; EMBL-European Bioinformatics Institute, Cambridge, United Kingdom; University of Warwick, Coventry, United Kingdom; NIHR Oxford Biomedical Research Centre, Oxford University Hospitals NHS Foundation Trust, John Radcliffe Hospital, Oxford, United Kingdom

**Keywords:** Carbapenemase-producing Enterobacterales, genomic epidemiology, bla_KPC_, wastewater sites, healthcare-associated

## Abstract

Healthcare-associated wastewater reservoirs and asymptomatic gastrointestinal patient colonisation by carbapenemase-producing Enterobacterales (CPE) contribute to nosocomial CPE dissemination. We systematically sampled wastewater sites (n=4488 sampling events; 349 sites) and patients (n=1247) across six wards over 6-12 months in 2016 to better understand bla_KPC_-associated CPE (KPC-E) diversity within these niches and transmission potential in an endemic healthcare setting. Up to five isolates in KPC-E-positive samples were sequenced (Illumina). Recombination-adjusted phylogenies were used to define genetically related strains; assembly and mapping-based typing approaches were used to characterise antimicrobial resistance genes, insertion sequences, and Tn4401 types/target site sequences. The wider accessory genome was evaluated in a subset of the largest clusters, and those crossing niches.

Wastewater site KPC-E-positivity was substantial (101/349 sites [28.9%] positive); 228/5,601 (4.1%) patients cultured were CPE culture-positive over the same timeframe. At a genomic-level, 13 KPC-E species and 109 strains were identified, and 24% of wastewater and 26% of patient KPC-E-positive samples harboured ≥1 strain. Most diversity was explained by the individual niche, suggesting localised factors are important in selection and spread. Tn4401+target site sequence diversity was greater in wastewater sites (p<0.001), which might favour Tn4401-associated transposition/evolution. Shower/bath and sluice/mop-associated sites were more likely to be KPC-E-positive (Adjusted Odds Ratio [95% CI]: 2.69 [1.44-5.01], p=0.0019 and 2.60 [1.04-6.52], p=0.0410, respectively). Different strains had different transmission and bla_KPC_ dissemination dynamics.

There may be substantial KPC-E colonisation of wastewater sites and patients in KPC-E-endemic healthcare settings. Niche-specific factors (e.g. microbial interactions, selection pressure) likely affect carbapenemase gene persistence and evolution, and different strains and mobile genetic elements with different transmission dynamics influence carbapenemase gene dissemination; these factors should be considered in surveillance and control strategies.

## 3. Impact statement

Antibiotic resistance is a major healthcare problem, and common infection-causing bacteria known as Enterobacterales have developed resistance to one of the “last-line” antibiotic classes used for treatment, namely carbapenems. Carbapenem resistance in Enterobacterales is largely caused by the presence of carbapenemase enzymes, such as “KPC” (Klebsiella pneumoniae carbapenemase), which has caused outbreaks of antibiotic-resistant healthcare-associated infection in the USA and UK. Many infection-associated Enterobacterales species can also live asymptomatically in the human gut and environmental niches, such as hospital wastewater sites, which act as potential reservoirs for the exchange of the gene encoding for KPC (bla_KPC_). Diversity within and transmission between these reservoirs remains poorly characterised. We therefore systematically and densely sampled humans and wastewater sites (e.g. sink/shower drains, toilets etc.) on six wards in 2016-2017 and sequenced 1646 isolates in a prospective study in a hospital affected by outbreaks of KPC-Enterobacterales-associated infections. We found that 1 in 20 patients and nearly a third of all wastewater sites were colonised with KPC-Enterobacterales. Samples frequently contained multiple different KPC-positive species or strains at any given time, and different transmission patterns were evident. This study has major implications for surveillance of carbapenem-resistant Enterobacterales in hospitals, and for designing strategies to mitigate their spread.

## 4. Data summary

The authors confirm all sequencing data and available metadata has been provided in NCBI BioProjects PRJNA768622 and PRJNA514245 and in supplementary dataset 2.

## 5. Introduction

Carbapenemase-producing Enterobacterales (CPE) are a global health threat^1^, and commonly associated with multi-drug resistance, rendering treatment of CPE infections difficult. Major transmissible global carbapenemases include the metallo-beta-lactamases (bla_NDM_, bla_VIM_, bla_IMP_), some oxacillinases (blaOXA_-48/48-like_ variants), and the Klebsiella pneumoniae carbapenemase (KPC, encoded by bla)^2^. Intra-and inter-species horizontal transfer of these genes is facilitated by mobile genetic elements such as transposons and plasmids, resulting in rapid dissemination of carbapenem resistance^3, 4^.

CPE have been isolated from human and animal gastrointestinal tracts, and from sewage, rivers and sink drains^5–7^. Recent studies have highlighted that hospital wastewater sites may act as a CPE reservoir^5^, but the diversity within these niches, genetic overlap with patient isolates and likely directionality and rates of transmission remain to be elucidated. Studies evaluating the contribution of patient-to-patient transmission as the sole explanatory factor explaining transmission have generally been unable to robustly explain the majority of transmission events, suggesting that patient, staff and/or environmental reservoirs remain insufficiently sampled to account for the diversity observed^8^. Few studies have considered within-niche diversity of strains by sampling multiple isolates per individual and/or site^9–11^.

The Manchester University NHS Foundation Trust (MFT; Manchester, UK), has experienced on-going transmission and patient acquisition of multi-species CPE, predominantly bla_KPC_-positive Enterobacterales (KPC-E), since 2009. Following a bla_KPC-2_-ST216-Escherichia coli outbreak on two cardiac wards in the Manchester Royal Infirmary (MRI; part of MFT) and subsequent ward closure and plumbing infrastructure replacement, early environmental sampling after the ward reopened suggested rapid re-colonisation of wastewater sites with KPC-E, likely from transfers of colonised patients to the ward and ongoing evolution of new KPC-E strains in the environment^12^. We therefore undertook a prospective study systematically sampling all wastewater sites on six wards for 6-12 months alongside patient rectal screening/clinical sampling, and used an anonymised electronic database to characterise patient admission, ward movement, sampling profiles and culture results. Multiple colonies were isolated from KPC-E-positive samples to define within-niche diversity, and whole genome sequencing (Illumina) of individual isolates was used to characterise this diversity and consider modes of evolution and transmission.

## 6. Methods

### Study setting and epidemiological data

MFT is a large, regional referral centre in northwest England, UK, managing >10,000 patients/year. In response to the regional emergence of KPC-E^13^, the Trust implemented an extensive Infection Prevention and Control (IPC) programme, consistent with UK CPE guidelines (published in 2014)^14^. Despite this, in April 2015 there was a large bla -E. coli outbreak in the cardiac unit^12^, leading to its closure (September 2015-January 2016) and complete refurbishment, including plumbing replacement. Subsequently systematic, prospective wastewater site sampling was undertaken (described further below).

To evaluate patient-level microbiological and admissions data, MFT electronic bacteriology records were linked, based on NHS numbers, to patient administration data, and anonymised. For this study, we analysed anonymised patient data from 1^st^ January 2016-31^st^ December 2016 inclusive, and included only those patients with exposure to any one of six wards within the three study units (acute medicine, cardiology, geratology) during that time.

### Environmental sampling and laboratory processing

All sink/drain/wastewater sites on six wards in three units were sampled: the cardiac unit (wards 3 [W3] and 4 [W4]); the geratology unit (wards 45 [W45] and 46 [W46]); and the acute medicine unit (wards AM1 and AM2) (Table S1 shows sampling site designations). All wastewater sites on both W3 and W4 were sampled fortnightly on rotation from 8^th^ January 2016-28^th^ December 2016. W45, W46, AM1, AM2 wastewater sites were similarly sampled fortnightly, but from 18^th^ July 2016-31^st^ December 2016. Wastewater site types are described in the Supplementary methods.

Environmental sampling was carried out by aspirating ∼20mls of wastewater from sink P-traps, shower drains or toilets and performing enrichment-based culture (see Supplementary methods). Multiplex, real-time, quantitative PCR (for bla, bla, bla ^12^) was performed on broths following incubation. 10μl of any bla_KPC_-positive-broth were streaked onto CPE-selective agar plates (Chromid CARBA) and re-incubated (aerobically, 37°C).

### Patient sampling and laboratory processing

Routinely collected patient clinical samples for the diagnosis of infection were processed using standard operating procedures in line with UK Standards^15^. In addition, a rectal CRE screening programme was in place from 2014 in line with national guidance: This included routine screening of high-risk patients (i.e. previously CRE-positive and/or a history of hospitalization abroad or in UK hospital with a known CRE problem in the last 12 months) on admission and weekly; screening of all patients on wards where CRE-positive patients were identified (either colonised or infected cases); ward closure and twice weekly screening if more than two cases were identified on a ward until no new cases were identified; isolation and/or cohorting of CRE-positive patients. Rectal swabs were screened for CPE using the Cepheid Xpert Carba-R assay or an in-house multiplex PCR (identifying bla_KPC_, bla_NDM_, and bla_OXA-48_). Species identification of isolates was performed by MALDI-ToF (Bruker Biotyper, Bruker MD); antimicrobial susceptibility testing was performed as per EUCAST guidelines^16^. Enterobacterales isolates that had resistant or intermediate susceptibility to any carbapenem were classified as a carbapenem-resistant Enterobacterales (CRE).

### Analyses of within-niche diversity

To evaluate within-niche diversity in the environment and patients, up to five different CRE colonies were individually sub-cultured; bla_KPC_-positivity in sub-cultures was re-confirmed by PCR and species identification performed by MALDI-ToF prior to storage in nutrient broth at -80°C.

### DNA extraction and isolate sequencing

DNA was extracted from frozen sub-culture stocks with the QuickGene kit, as per the manufacturer’s instructions, with an additional mechanical lysis step post-chemical lysis (6m/s at 40secs x2 FastPrep, MPBio). Isolates were sequenced on the Illumina HiSeq 4000 generating 150bp paired-end reads.

### Sequence data processing and analysis

Sequencing reads were initially processed using an in-house pipeline^17^ (details in Supplementary methods); species identification was performed with Kraken2 (default settings)^18^. If at least 50 sequenced isolates were of a single species, reads were mapped to species-specific references. Variant calling and filtering was performed, and consensus fasta sequences of variant calls with respect to the reference generated. Recombination-adjusted phylogenies by species were created from core chromosomal single nucleotide variants (SNVs; padded to the length of the reference genome prior to analysis) using IQtree^19^ (flags: -m GTR+G -blmin 0.00000001 -t PARS) and then ClonalFrameML^20^ (default parameters). A distance of ≤400 SNVs was used as a threshold to define clusters based on previous work (threshold used similar to^21^; distribution of SNV distances within clusters displayed in Fig.S2A-E for the most common species identified); indels were not considered in our evaluation of relatedness. For detailed sub-strain analysis for K. pneumoniae strain 9 (see Results), sub-strains were defined on the basis of accessory genome clusters characterised using Panaroo (default parameters including --clean-mode strict^22^) and clustered using the R heatmap package.

Isolates were assembled using SPAdes (v3.6; default parameters)^23^. Multi-locus sequence types were derived using BLASTn and species-specific databases (https://pubmlst.org; Achtman 7-gene MLST for E. coli). AMR genes were identified using ARIBA^24^ with the CARD database^25^ and insertion sequences using ISFinder (https://isfinder.biotoul.fr). bla_KPC_ genes, Tn4401 transposon contexts and the 5-bp target site sequences (TSSs) on either side of the Tn4401 transposon reflecting Tn4401 transposition signatures were characterised using TETyper^26^, a Tn4401b reference sequence, a 5bp flank length and the default structural/SNV profiles. Arbitrary profile numbers were assigned based on the presence of unique profiles of AMR genes, plasmid replicons, IS sequences (see Supplementary dataset 1 for details of which AMR gene variants constitute a given AMR gene profile, and Supplementary dataset 2 for details on plasmid replicon and IS combinations identified).

For the analysis of K. pneumoniae strain 9 (see Results), putative transmission networks were inferred first using SCOTTI^27^ (Supplementary methods), and then heuristically, linking isolates identified as clusters on the basis of accessory component profiles and geographic/temporal overlap (Supplementary methods).

### Statistical analyses and data visualisation

Descriptive statistics were calculated in R v3.6.2 or STATA v16.1. To conduct permutational analyses of variance, we used the adonis function (vegan R package, v2.5-7)^28^ on a matrix of pairwise Gower distances based on genetic features (i.e. composite of species-strain type, plasmid replicon profile, AMR gene profile, IS profile, and Tn4401+flanking TSS profiles), which was calculated using the daisy function (R cluster package). To evaluate the impact of unit location or environmental site type on the odds of CPE positivity, we used logistic regression with robust standard errors clustered by environmental site sampled (R rms package, v6.2-0)^29^. Data visualisation was using the ggplot2^30^ and scatterpie^31^ (v.0.1.5) R packages, and BioRender (www.biorender.com).

### Data availability

Sequence data have been deposited in NCBI’s GenBank (BioProject accession PRJNA768622 and PRJNA514245; accession numbers and metadata for successfully sequenced isolates are in Supplementary dataset 2). We are not able to share the complete database of patient tests, results and admissions episodes; dates for patient admission/sampling are given as month-year only. Of note, labels represented in the figures/supplementary data are anonymised study-specific labels that are not patient-identifiable.

## 7. Results

### Environmental sampling revealed high prevalence and clustering of wastewater site KPC-E colonisation

349 sites across the six study wards (i.e. three units) were sampled a total of 4,488 times (Fig.1): 101/349 (28.9%) sites and 319/4,488 (7.1%) sampling events were KPC-E-positive, with no difference by unit (Fig.2, p=0.908), or over time. Adjusting for environmental site type and unit location, shower/bath drain sites and sluices/sluice sinks/mop sinks were significantly more likely to be positive for KPC-E than others (Adjusted Odds Ratio [AOR] (95% CI): 2.69 (1.44-5.01), p=0.0019 and 2.60 (1.04-6.52), p=0.041, respectively), and toilet water sampling less likely to be positive (AOR (95% CI): 0.28 (0.10-0.78), p=0.015); there was no evidence of a unit-dependent difference (Table 1). 7/349 (2.0%) sites were persistently KPC-E positive (≥10 consecutive KPC-E-positive sampling events); interestingly five of these were medicines/treatment room handwash basin drains (of eight medicines/treatment room sinks in total) likely only accessible to healthcare staff (Table S1, Fig.S3).

**Figure 1.**
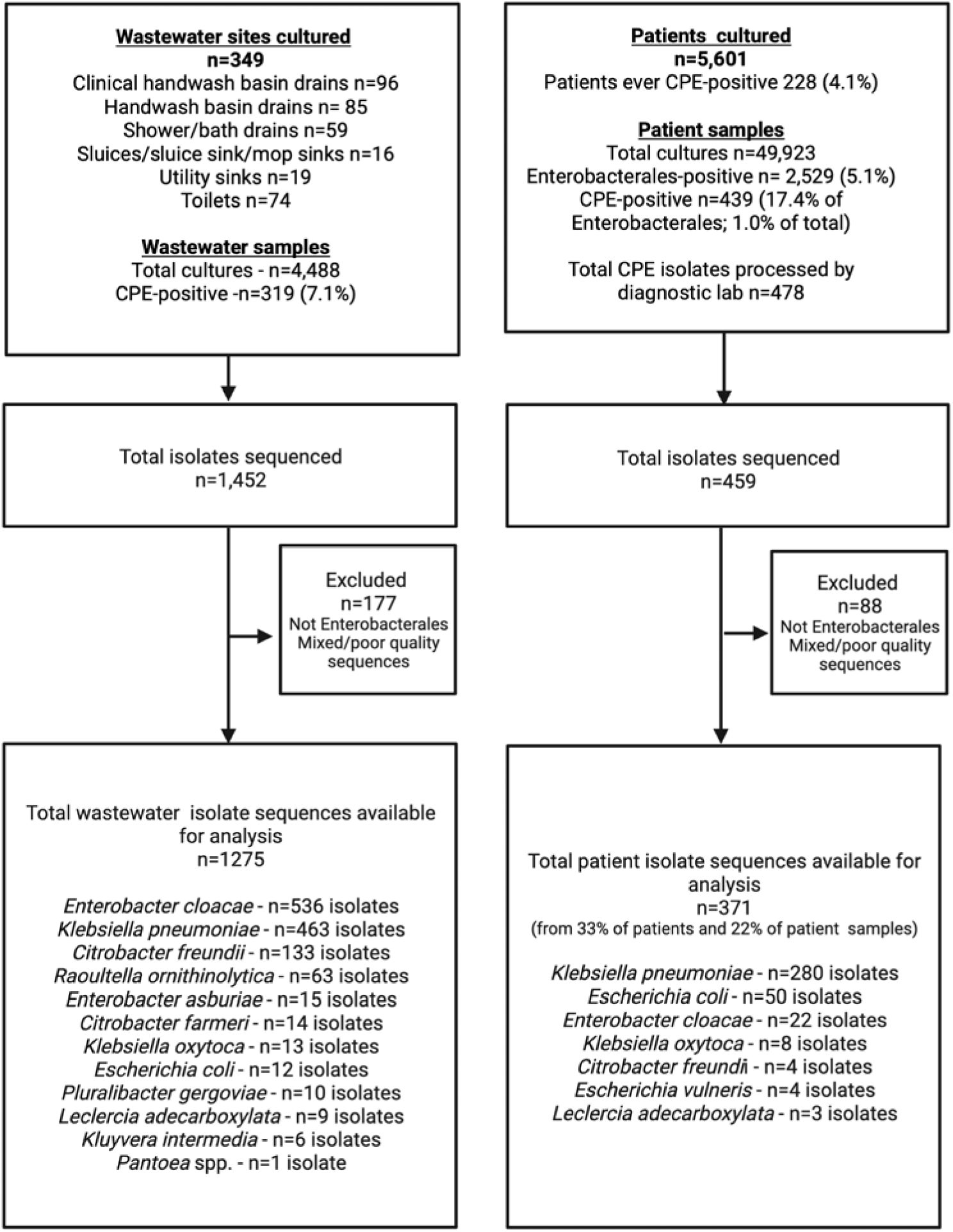
Sampling and sequencing flowchart for the study.

**Figure 2.**
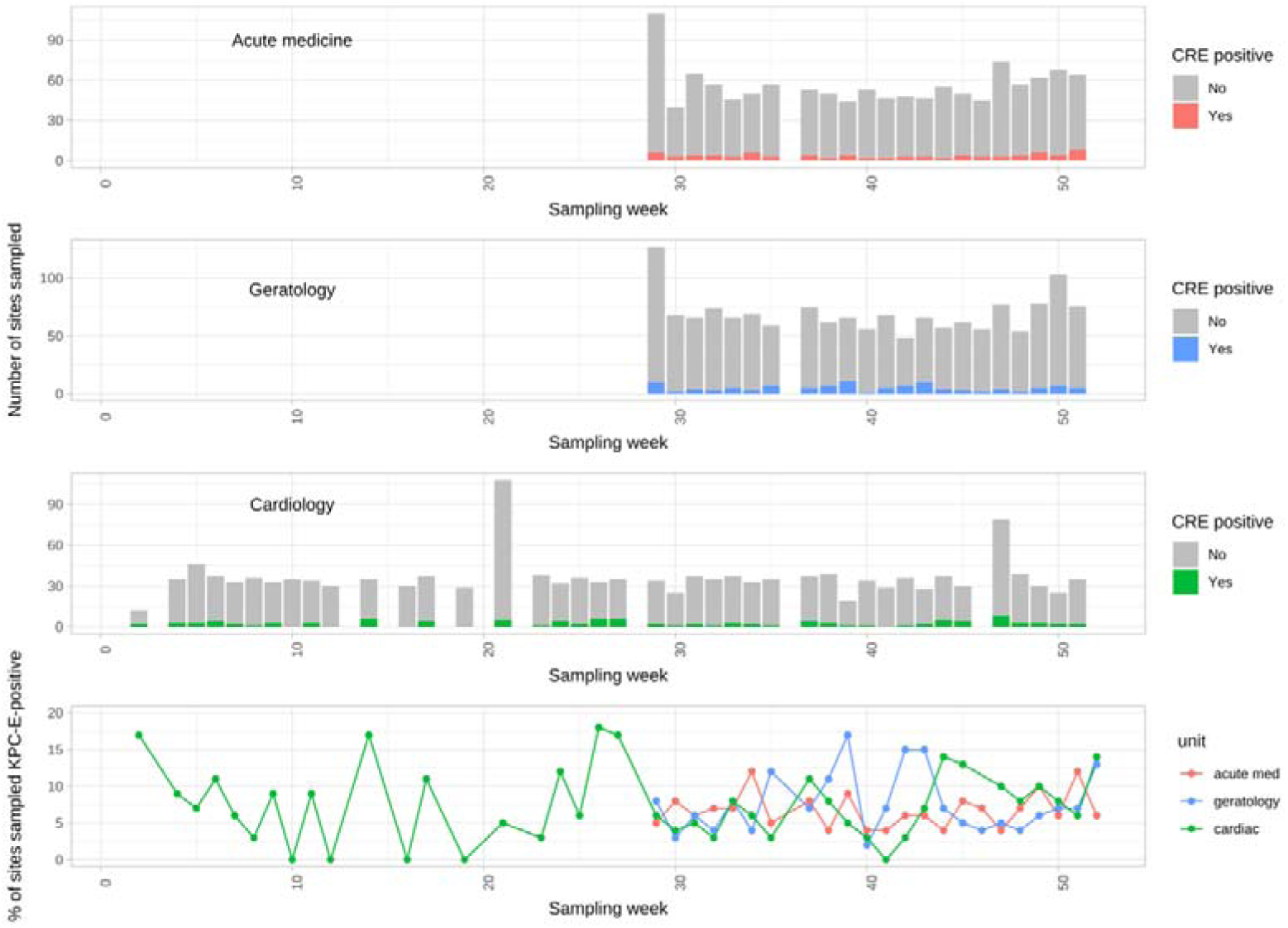
blaKPC-Enterobacterales (KPC-E)-positive environmental sites by week in 2016 and hospital unit. The number of environmental site samples performed and KPC-E-positive samples by week of sampling, stratified by unit (top three panels), and the proportion of KPC-E-positive sites by week of sampling, again stratified by unit (bottom panel).

**Table 1.** Association of environmental site type and unit location with bla_KPC_-Enterobacterales (KPC-E) positivity in environmental samples. Outputs of a multivariable logistic regression model with robust standard errors clustered by environmental site type and unit location; no other variables were considered in the model. P-values <0.05 were considered significant and are highlighted in bold. No interactions with an interaction Wald p <0.05 were observed.

| Environmental site | Number of sites | Number of sampling events KPC-E-positive (% of total sampling events) | Number of sampling events KPC-E-negative | OR (95% CI) | p |
| --- | --- | --- | --- | --- | --- |
| **Environmental site type** |  |  |  |  |  |
| Clinical handwash basin drain | 96 | 79 (5.9%) | 1270 | Reference |  |
| Handwash basin drain | 85 | 52 (4.6%) | 1077 | 0.99 (0.51-1.93) | 0.9735 |
| Shower/bath drain | 59 | 100 (19.2%) | 421 | 2.69 (1.44-5.01) | **0.0019** |
| Sluice/sluice sink/mop sink | 16 | 47 (17.9%) | 215 | 2.60 (1.04-6.52) | **0.0410** |
| Toilet | 74 | 5 (0.5%) | 931 | 0.28 (0.10-0.78) | **0.0152** |
| Utility sink | 19 | 36 (12.4%) | 255 | 1.32 (0.47-3.68) | 0.5965 |
| **Environmental site location** |  |  |  |  |  |
| Acute medicine unit | 129 | 86 (6.6%) | 1223 | Reference |  |
| Cardiac unit | 76 | 111 (7.0%) | 1453 | 1.07 (0.57-2.00) | 0.8412 |
| Geratology unit | 144 | 122 (7.6%) | 1493 | 0.88 (0.52-1.48) | 0.6243 |

### Approximately one in twenty-five patients was affected by carbapenemase-producing Enterobacterales (CPE), with most cases reflecting rectal colonisation with KPC-producing K. pneumoniae, E. coli or E. cloacae

Of 2,529 Enterobacterales positive-samples (2,622 Enterobacterales isolates, 1,247 patients), 439/2,529 (17.4%) samples cultured at least one carbapenem-resistant Enterobacterales (CRE, n=478 isolates in total) (Figs.1, S4). For 344/439 (78.4%) CRE-positive samples on which carbapenemase PCR data were also available, all contained at least one known carbapenemase gene, of which 303/344 (88.0%) were bla_KPC_ (Table S2, CRE therefore henceforth denoted as CPE for simplification). CPEs were cultured from 228 patients (4.1% of all patients cultured, 18.2% of all patients with an Enterobacterales-positive culture) during the study period (median=1, IQR: 1-2, range: 1-17 CPE isolates/patient).

The majority of CPE-positive isolates came from rectal screens (n=387/478 [81.0%]); others came from non-screening clinical specimens (91/478 [19.0%]) (Table S3). Most CPE-positive patients (178/228 [78%]) were CPE-positive only on rectal screen, but 23/228 (10%) individuals were positive only on clinical culture, and 27 (12%) on both. By routine laboratory species identification methods, >20 species and eight genera were represented amongst CPEs, with K. pneumoniae, E. coli and E. cloacae complex predominating (434/478 [90.8%]). Notably, the proportions of carbapenem-resistant isolates within these top three carbapenem resistance-associated species varied (n=284/593 [48%] vs 106/1,404 [8%] vs 44/110 [40%] respectively; Fisher’s exact test, p<0.001, Fig.S4).

The proportion of patient CPE-positive cultures was lower than that of environmental site samples, typically ∼3% (Fig.3, bottom panel). Occasional peaks in prevalence were observed, consistent with outbreaks (e.g. week 31, acute medicine unit). CPE prevalence amongst patient cultures on the refurbished cardiology unit was 0% (97.5% CI: 0-0.02) for the first 10 weeks post-reopening, although two utility room sink drains on were positive for KPC-E a day after W3 reopened to patient admissions (11^th^ January 2016) following closure and complete plumbing replacement over three months; thereafter the unit was rapidly re-colonised (Fig.2).

**Figure 3.**
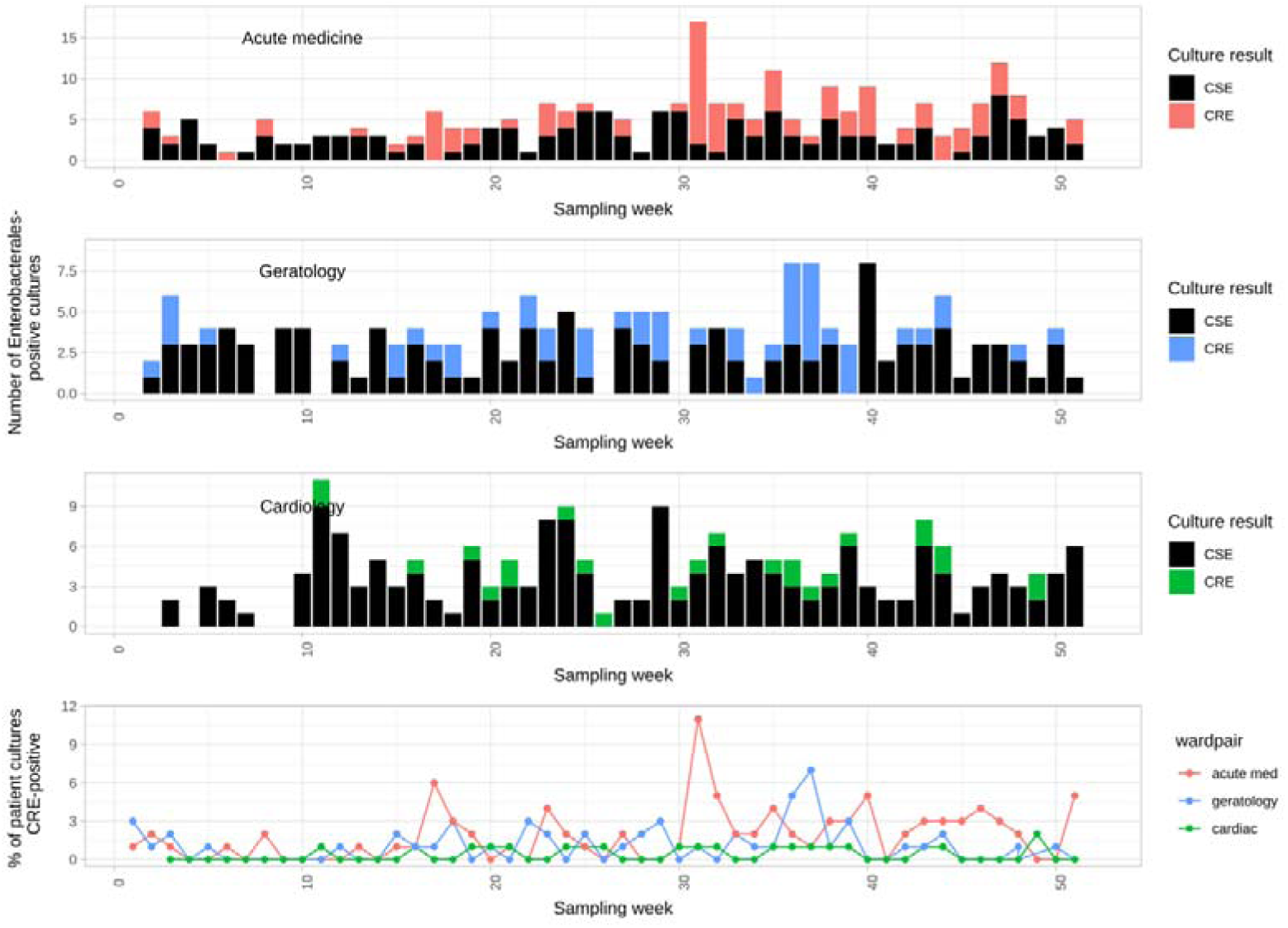
Carbapenem-susceptible Enterobacterales (CSE) and carbapenem-resistant-Enterobacterales (CRE) patient cultures by week in 2016 and hospital unit. Top three panels show counts of Enterobacterales culture-positive samples stratified by unit and carbapenem susceptibility; bottom panel shows the proportion of all cultures taken that were CRE, again stratified by unit.

### Structuring of diversity by patient and environmental niche, with most diversity explained by the individual niche

1,646 CPE isolates were successfully sequenced (1,275 environmental, 371 patient isolates; Fig.1, Supplementary dataset 2), of which all contained bla_KPC-2_ as the carbapenem resistance variant, and 13 bla_KPC-2_-Enterobacterales species were identified across patients and environmental reservoirs. Amongst five species with ≥50 sequenced isolates which were evaluated at the strain-level, 109 unique strains were represented. Of the 109 strains, 61 (56%) were found only in the environment, 27 (24%) only in patients and 21 (19%) in both patients and the environment (Fig.4 for the major species); the median strain cluster size was 5 isolates (range: 1-122), and the median number of unique niches (i.e. unique patients or wastewater sites) affected by a strain was 2 (range: 1-23).

**Figure 4.**
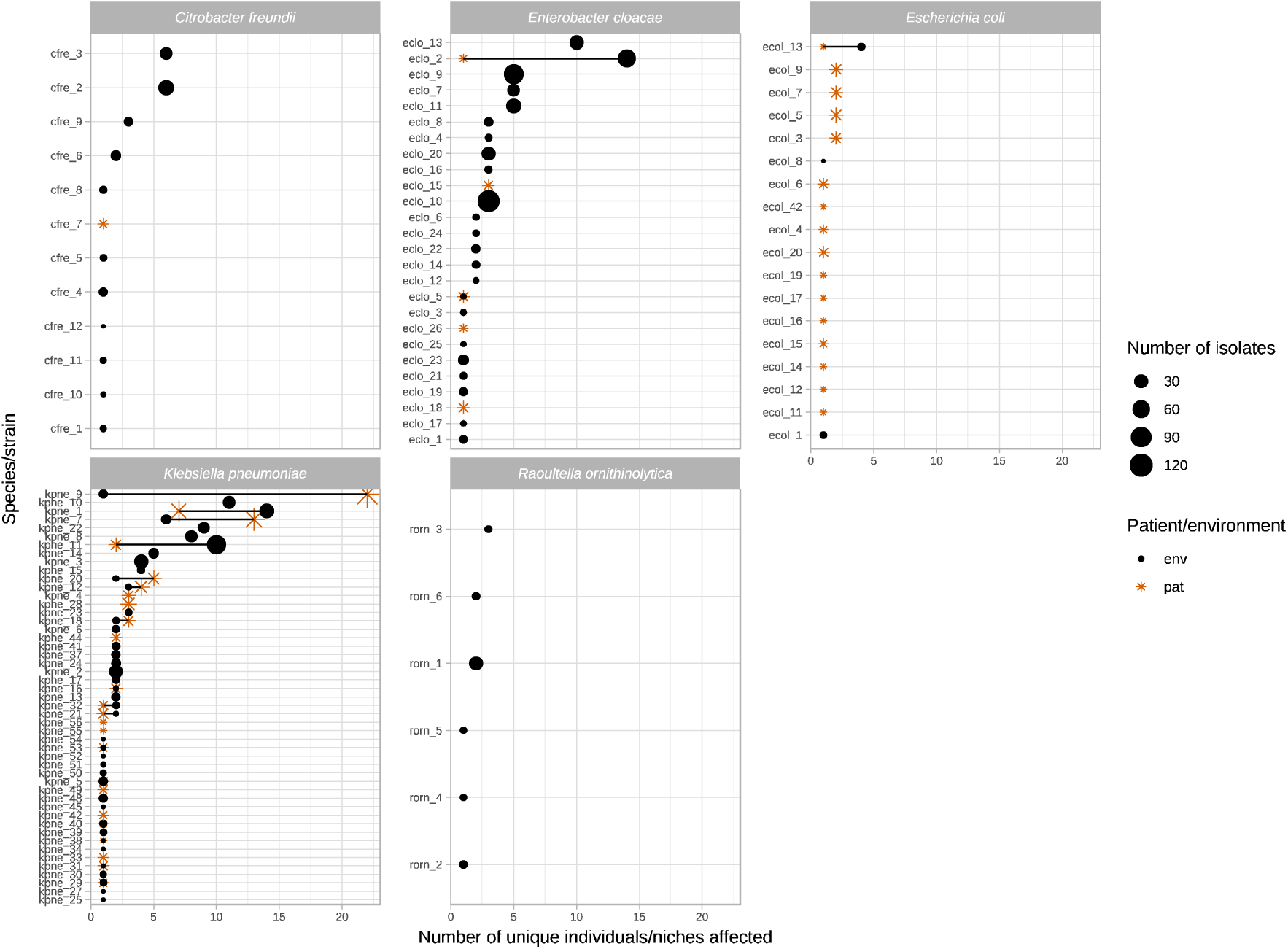
Number of unique patient/environmental niches colonised by common blaKPC-2 Enterobacterales species/strains and the total number of isolates for each cluster. Includes species/strains if >50 isolates of a species were identified [see Methods]). Orange stars denote patient niches, black circles wastewater niches; larger shape size denotes a larger number of isolates sequenced. Strains observed in both niches are either overlapping shapes (same number of niches), or joined by a line (different numbers of niches).

Amongst sequenced isolates, 100 plasmid replicon profiles, 754 IS profiles, 939 AMR gene profiles, and 70 Tn4401/TSS types were identified (Supplementary dataset 2). A much greater proportion of all identified Tn4401/TSS types was observed in the environment versus patients (66/70 [94%] of types versus 12/70 [17%] respectively; Fisher’s exact test, p<0.001), and all the isolates with >1 set of TSSs were seen in environmental sites, consistent with these representing a more favourable niche for Tn4401-associated transposition/evolution and dissemination.

For colonised niches (i.e. patient or environmental), using a metric of genomic diversity based on strain, plasmid replicon profile, AMR gene profile, IS profile, and Tn4401+5bp TSS profiles, most variance was explained by the specific individual site sampled (R^2^=50.5%; p<0.001), versus small but significant contributions made by niche type (i.e. patient vs environmental niche; R^2^=5.6%; p<0.001) and unit location (R^2^=2.8%; p<0.001).

For the fifteen scenarios where exactly the same strain, AMR gene profile, IS profile, plasmid profile and Tn4401/TSS type were seen in patient and environmental niches (Fig.S5), and considering only the event-pairs where the chromosomal SNP distances were within 5 SNPs of each other and there was a change in sampling site, the temporal relationship of these supported: patient-to-environment transmission in 13/29 (45%) events, environment-to-patient transmission in 2/29 (7%) events, environment-to-environment transmission in 1/29 (3%) events, and patient-to-patient transmission in 13/29 (45%) events (test of equality of proportions, p<0.001).

### Environmental and patient niches harbour diverse KPC-E within single samples and over time

Environmental sites showed evidence of major strain-level diversity: from the 101/349 sites from which bla_KPC-2_-Enterobacterales isolates were successfully sequenced at any timepoint, 78/319 (24.4%) KPC-E-positive samples had >1 bla_KPC-2_-Enterobacterales strain, namely 52/319 (16.3%) with two strains, 16/319 (5.0%) with three, 9/319 (2.8%) with four and 1/319 (0.3%) with five strains (Figs.5, S6). Over the study period, colonised environmental sites had a median of 3 (range: 1-17, IQR: 2-5) bla_KPC-2_-Enterobacterales strains. 49/101 sites (49%) were apparently colonised by a single bla_KPC-2_-Enterobacterales strain only, of which 38 were at single sampling-points, and 11 sites positive with the same strain at ≥2 timepoints.

For patient samples, based on routine laboratory microbiology, evidence of mixed-species KPC-E colonisation/infection was found in 35/439 (8%) samples from 26/228 (11%) patients; these were mostly E. coli/K. pneumoniae mixtures (n=17 samples; maximum two species identified). To evaluate within-patient diversity with genomic rather than species-level resolution, in a subset of 76 patients, strains were sequenced successfully from at least one CPE-positive sample during the study period; for 97 samples, multiple colonies were picked from single sample sub-cultures. In total 457 sequenced colonies from 97 samples were available for evaluation of within-patient diversity; 25/97 samples (25.7%) had more than one strain per sample, namely 17/97 samples (17.5%) with two bla_KPC-2_-Enterobacterales strains, 6/97 (6.2%) with three strains and 2/97 (2.0%) with four strains (Figs.5, S6) - i.e. considerably more diversity identified than based on routine laboratory species identification data (26% vs 8% samples; Fisher’s exact test, p<0.001).

For a subset of 14 patients and 53 wastewater sites where within-sample diversity was evaluated by sequencing multiple isolates from individual samples and ≥2 longitudinal samples were positive, longitudinal switches in bla_KPC-2_-Enterobacterales species-strain composition was common, occurring in 7/14 (50%) of patients and 41/53 (77%) wastewater sites (Figs.5, S6).

### Genomic analysis reveals distinct population biology for different bla_KPC-2-_Enterobacterales strains

To investigate different modes of bla_KPC-2_ dissemination, we characterised in detail several sequenced KPC-E strain clusters, including the largest environmental-only cluster (E. cloacae strain 10 [n=122 isolates]), a large cluster involving patients and the environment (K. pneumoniae strain 9 [n=106 isolates], and a cluster in which signatures of Tn4401 transposition appeared highly frequent (K. pneumoniae strain 11 [n=96 isolates]).

### Scenario 1: Dynamic changes in AMR gene, IS, plasmid replicon and Tn4401 profiles in E. cloacae strain 10 in sink drain sites

E. cloacae strain 10 (eclo10, ST32) appeared a site-restricted but persistent coloniser of three co-located sink drains on a geratology ward (in a drug room clinical handwash basin drain [F45], utility sink drain [F46], and a separate treatment room clinical handwash basin drain [F47]) over ∼5 months (18/Jul/2016-20/Dec/2016). At the core chromosomal level, strains appeared largely structured by site, with 7 SNVs separating the majority of isolates in F45 from those in F47, and 14 SNVs those in F46 from F45. Three isolates in F47 and one isolate in F46 clustered with the F45-associated isolates; sampling dates suggest that these represented transmission events from F45 (Fig.6A). There was evidence of rapid churn in AMR gene and IS content (111 and 36 different profiles respectively; Fig.6Bi and 6Bii) in the relatively stable strain background, and almost all isolates in F47 had acquired an additional set of IncHI2/HI2A replicons on a background of FIB/FII, consistent with plasmid gain/loss (Fig.6C). Although the Tn4401 type and flanking sequences were largely identical (Tn4401a-ATTGA-ATTGA in 105/122 (86%) isolates across sinks), there was evidence of unique additional Tn4401 deletions occurring in multiple isolates at several distinct timepoints in the three sink sites (Fig.6).

**Figure 5.**
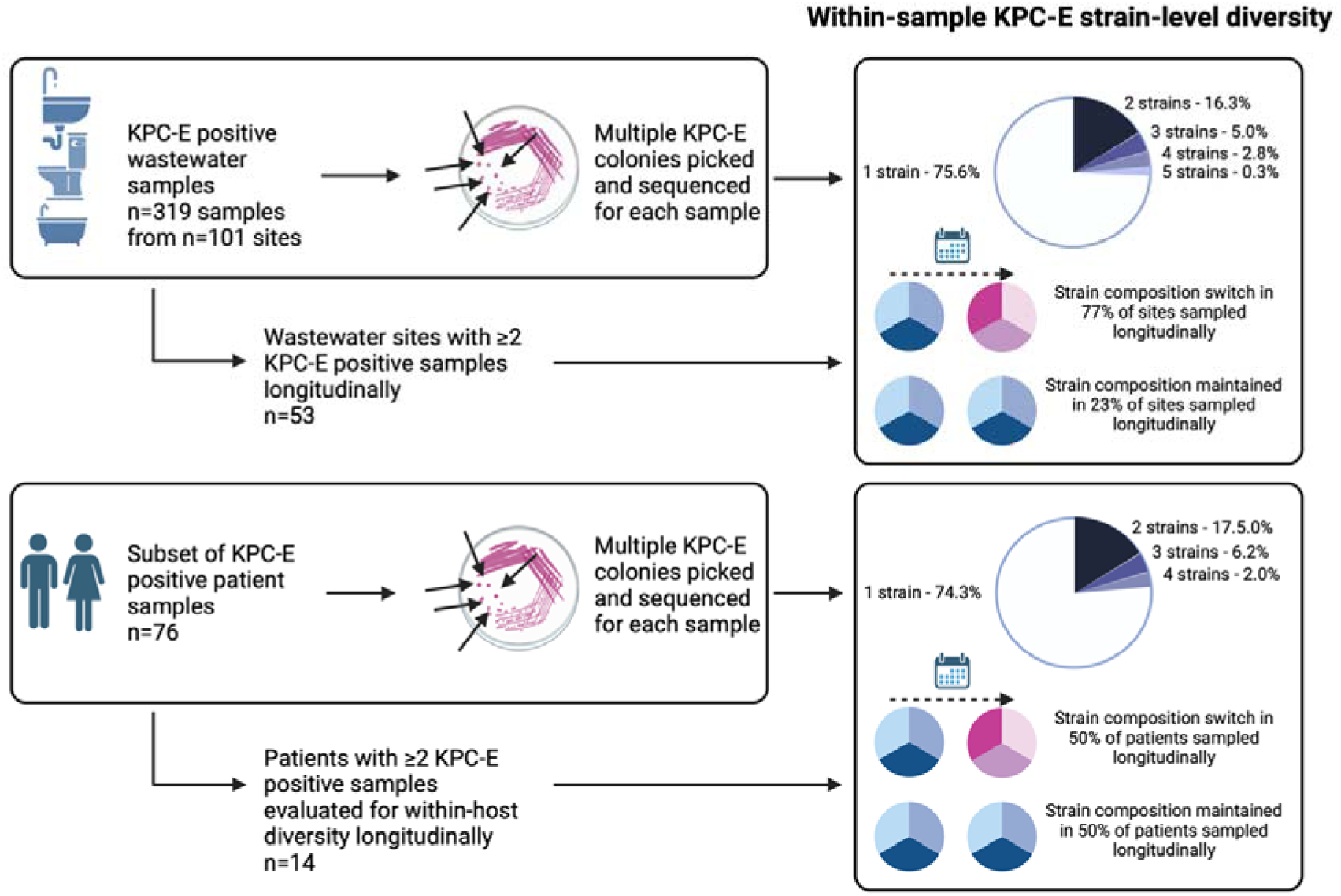
Summary of within-niche and KPC-E sample diversity using genomics.

**Figure 6.**
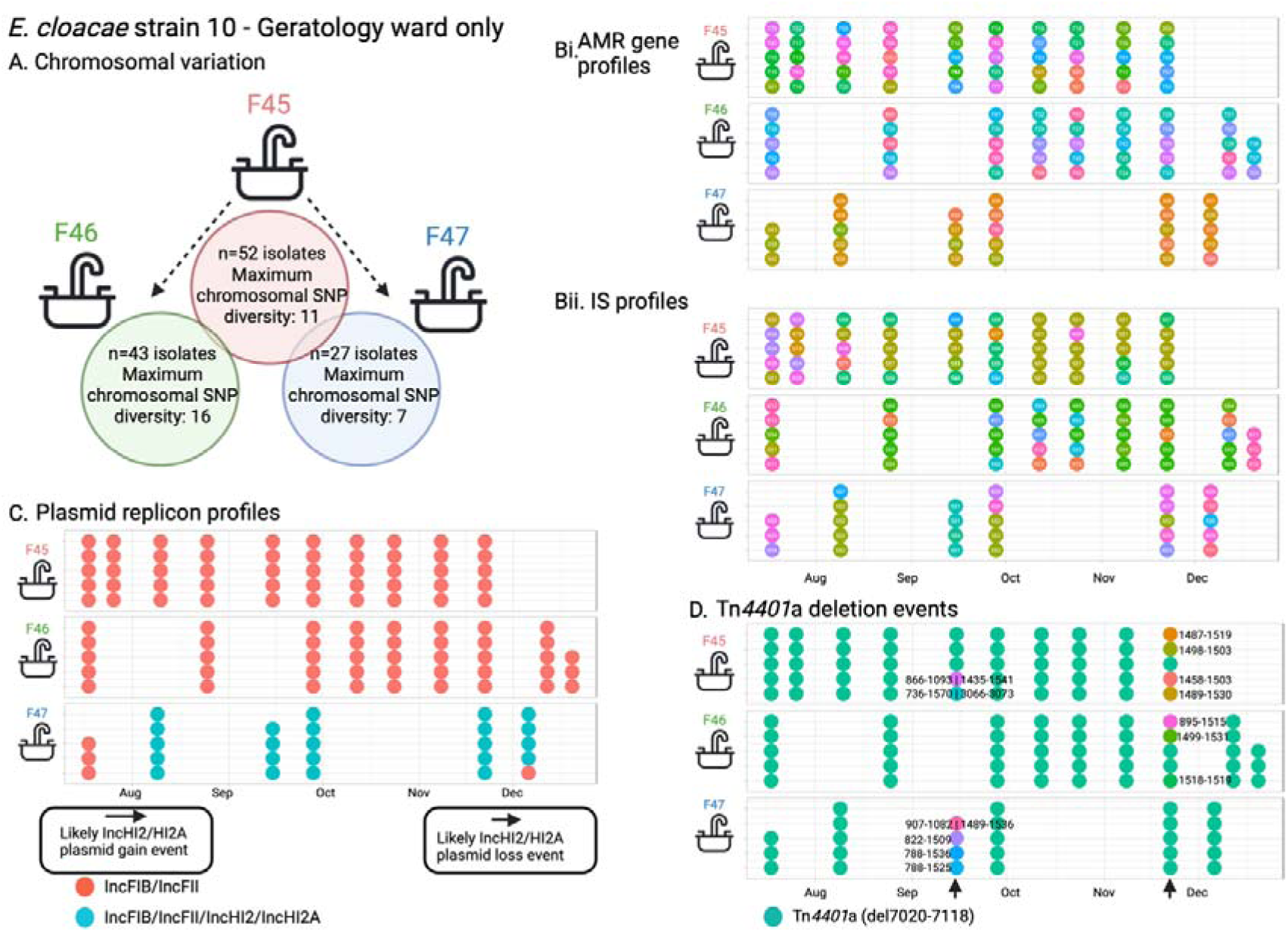
Enterobacter cloacae strain 10 - features of population structure and transmission dynamics - niche restriction, persistence and genetic turnover. Summaries of A) Chromosomal SNP distances for E. cloacae strain 10 across the three sites it colonised; Bi) AMR and Bii) IS profiles, C) plasmid replicon profiles, and D) deletions in Tn4401a in isolates with each sequenced isolate plotted as a dot reflecting within-sample diversity at given timepoints and longitudinally.

### Scenario 2: Rapid clonal patient-patient transmission of K. pneumoniae strain 9

K. pneumoniae strain 9 (kpne9, ST252) was highly related (Fig.S7; maximum 6 SNVs across strain, 68 isolates with 0 SNVs between them), and spread amongst 21 patients across three wards on two units (acute medicine AM1 and AM2 and geratology W46), with a dense outbreak in Aug 2016, most consistent, at least initially, with direct/indirect patient-patient transmission (Fig.7). bla_KPC_ was consistently nested in Tn4401-ATTGA-ATTGA, except for one isolate which acquired a Tn4401-associated mutation (isolate: 2216698_14, C4620T), and another in which the right target site sequence (TSS) could not be identified (isolate: 2035791_11). Amongst the 106 isolates there was evidence of four transient plasmid replicon acquisitions on a background of the stable presence of a set of IncFIB/IncFII replicons (Fig.S7). Transmission probabilities inferred by SCOTTI were weak (see Supplementary material; Fig.S8), as seen in other studies^8^, but using accessory cluster profiling to further discriminate amongst strains suggested that there were several discrete transmission clusters (Fig.S9), with some individuals involved in several of these (i.e. harbouring multiple distinct sub-lineages simultaneously, Fig.7). A single shower drain site was involved (A13), which was likely a seeded bystander from affected patients, but may have contributed to transmission of sub-strain 3g (Fig.7).

**Figure 7.**
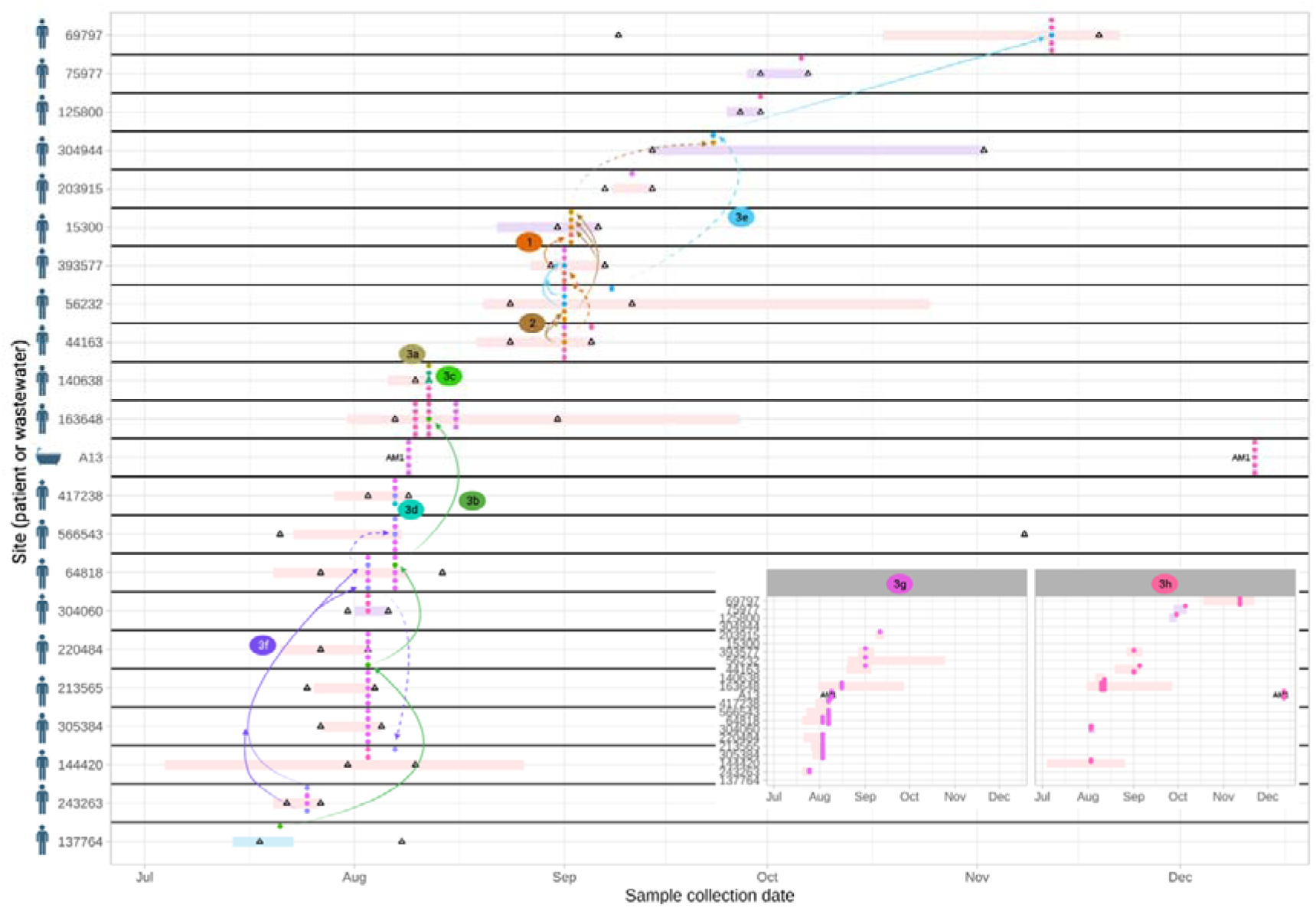
Klebsiella pneumoniae strain 9 - features of population structure and transmission dynamics - rapid patient-patient transmission. Putative transmission network for sub-lineages of K. pneumoniae strain 9, defined by accessory genome clustering (clusters 1, 2, 3a-h). Horizontal bars represent admission episodes to study wards for each patient (pale blue=W45, geratology; pale pink=AM1, acute medicine; pale purple=AM2, acute medicine; A13 represents the only environmental (shower) site involved. Empty black triangles denote CPE-negative rectal screens on patients. Arrows denote possible transmission links (dashed arrows denote events that have no geographic overlap or could be associated with two links based on negative screen, geographic location and timing). For clarity, arrows are not drawn for clusters 3g and 3h, which affected the most patients - these are shown in detail as an inset panel at the bottom right. In some cases patients were colonised by other species-strains contemporaneously (not shown here).

### Scenario 3: Cross-unit dissemination with evidence of significant Tn4401-associated bla_KPC_ transposition in K. pneumoniae strain 11

Given that Tn4401 transposition is associated with bla_KPC_ dissemination^21^, we also evaluated whether the number of different Tn4401-TSS types was different across strains, noting that the observed frequency of different Tn4401-TSS combinations was higher in K. pneumoniae strain 11 (kpne11, ST11) than in other strains (Fig.S10). On further analysis, Kpne11 represented two sub-clusters, separated by ∼40SNVs (Fig.8, designated “cluster 1” and “cluster 2”; Fig.S11), both of which were isolated from patients and wastewater sites across ward settings, including the cardiac unit after plumbing replacement. Within both clusters of isolates there were ten different Tn4401a flanking signatures observed, only one of which was seen in the other >1600 isolates sequenced in the study (Tn4401a-1_ATTGA_ATTGA, the most common flanking signature observed overall), and therefore most strongly consistent with the occurrence of an atypical and increased number of transposition events within this K. pneumoniae strain. Although all isolates contained IncFIB and IncFII plasmid replicons, evidence of plasmid gain/loss events as demonstrated by changes in plasmid replicon profiles also appeared common (Fig.8 for cluster 1).

**Figure 8.**
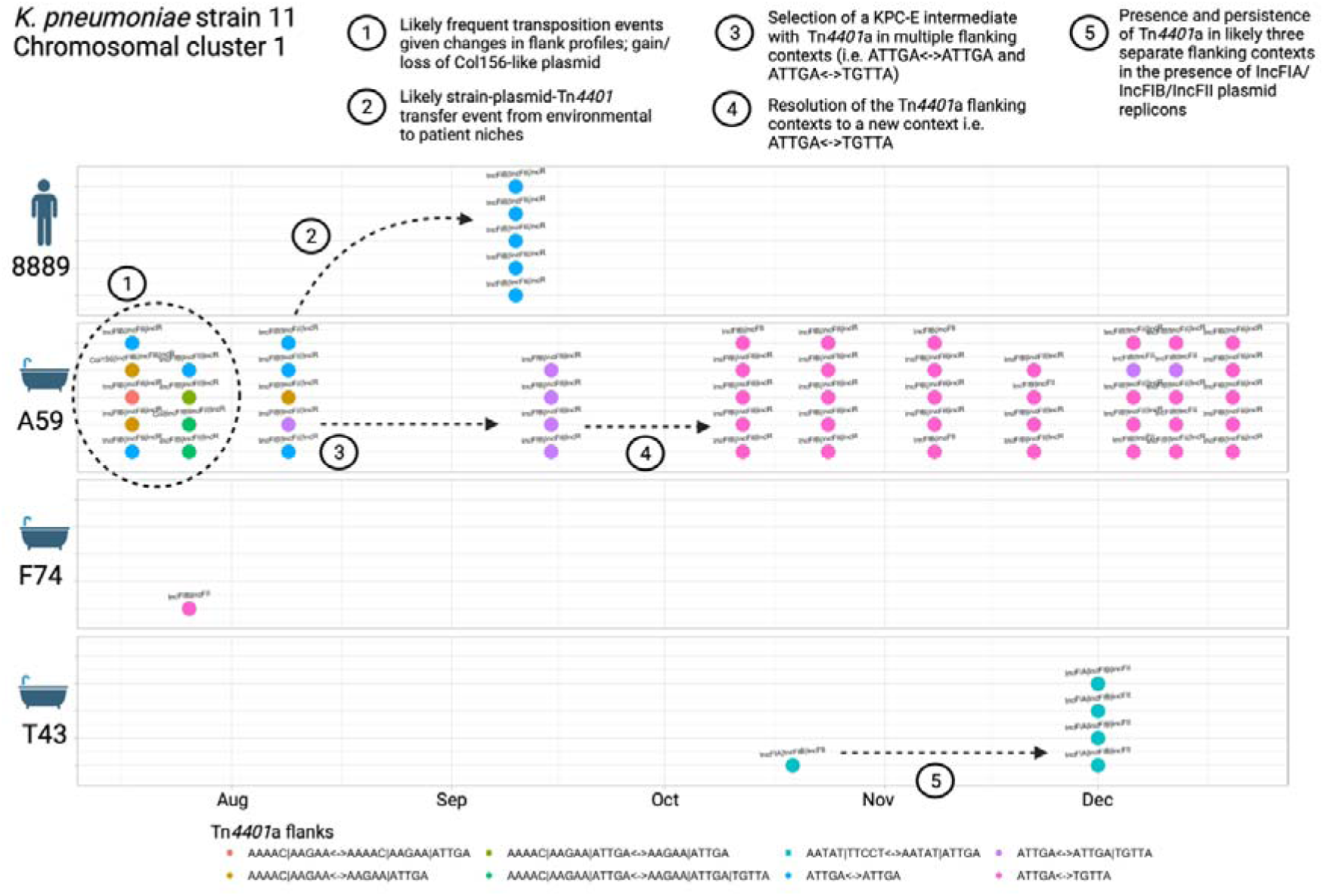
Klebsiella pneumoniae strain 11 cluster - features of population structure and transmission dynamics – multiple Tn4401 transposition events. Examples of possible transmission/genetic events are annotated on the figure. Each isolate sequenced is represented as a dot, with colour denoting the TSS profiles observed. Plasmid replicon profiles are annotated as text.

## 9. Discussion

This work highlights the substantial KPC-E diversity and flux that can occur in both human and wastewater niches in healthcare settings over variable timeframes, in this case in relation to a large bla_KPC-2_-associated carbapenemase gene outbreak. KPC-E strains have features likely to make generalising results regarding modes of transmission, evolution and gene sharing of bla_KPC_ amongst these species very challenging. Although structuring at the strain-level was observed by niche and species (Fig.4), likely transmissions occurred in a non-negligible manner in all directions from patients to other patients and the environment, and from the environment to patients and other environmental sites. Environmental niches and certain lineage contexts appeared to amplify bla_KPC-2_ transposition events.

Although some environmental sites were persistently colonised with individual strains over months, many appeared to be only transiently colonised. Sites particularly associated with KPC-E colonisation included sluices, sluice sinks, mop sinks and shower/bath drains, supported by other studies^5^, and consistent with these representing a major interface between human faeces and premise plumbing. Toilets were less likely to be KPC-E-positive, perhaps because of regular flushing or particular cleaning protocols; consistent with this, several studies have demonstrated that daily bleach application decontaminated wastewater sites^32, 33^.

Our study underscores the detailed and dense sampling effort required to understand within-sample diversity and longitudinal flux, and highlights the difficulty in accurately tracking the dissemination of AMR genes and strains across niches. This problem has also been demonstrated in other recent work investigating bla_OXA-48_, where only patient-based sampling was undertaken and most bla_OXA-48_-associated transmission events were non-delineated or only weakly supported by joint phylogenetic/epidemiological inference^8^. Our study clearly shows that different species and lineages represent different epidemiological risks and have their own behaviours, and a focus on single, clonal strains as mediators of AMR gene transmission is irrelevant in this and potentially many other settings where important AMR genes have become endemic.

The importance of evaluating asymptomatic carriage, characterising within-host diversity of strains, considering rapid acquisition/loss of strains over short timeframes, and including environmental sites in transmission networks has been partly demonstrated in previous studies investigating ESBL-E. coli^10, 34^ and Enterococcus spp.^35^, but generally these features are not jointly investigated because doing so is highly resource-intensive, and there are no robust analysis methods for doing this to our knowledge. However, without this effort, such as in studies which consider only single or infrequent sampling timepoints, and do not investigate asymptomatic colonisation or consider within-site diversity^36^, our study highlights that relevant events may easily be missed.

There are several limitations to this work. CPE-sample capture for patients for this study was not continuous, and we therefore only evaluated a subset of CPE-positive patients and samples using genomics (33% and 22% respectively); we anticipate many transmission events were missed, but our sampling already reveals the high complexity of bla_KPC_ transmission. We were not resourced to perform long-read sequencing to enable plasmid sequence reconstruction, and plasmid replicon or bla -contig profiling is relatively low-resolution and can be misleading^21^; we therefore avoided a detailed plasmid analysis which would be important in quantifying horizontal bla_KPC_ transmission. However, the diversity we observed in genetic contexts supporting bla_KPC_ was substantial, consistent with high rates of HGT. A further limitation is the absence of metadata capturing relevant selection pressures in each niche, including cleaning protocols for environmental reservoirs and drug prescription/antimicrobial stewardship data for patients. We only investigated one healthcare setting, but the same polyspecies, polyclonal phenomenon has been seen elsewhere and with other carbapenemase genes^4,8^. We focused only on Enterobacterales and may have missed wider HGT of carbapenemase genes to other bacteria. In line with this, at least 23 Pseudomonas spp., five Stenotrophomonas maltophilia and two Acinetobacter spp. that were PCR-positive for bla_KPC_ or bla_NDM_ were inadvertently picked up on rectal screening. Our approaches to evaluating some of the putative genetic changes and transmission events observed were heuristic and manual, partly because the data were highly hetereogenous and complex. The study is now relatively historic, but these types of polyspecies, polyclonal carbapenemase gene outbreaks associated with hospital wastewater sites continue to be described and as such we believe the findings remain of interest.

Further optimisation of methods to evaluate multi-level genetic transmission and within-sample diversity without individual colony-level characterisation is needed. Holistic approaches using fully reconstructed chromosome/plasmid assemblies to quantify the diverse types of genomic transmission (including clonal transmission, horizontal transfer at the gene, transposon and plasmid-level) are also required to better understand what facilitates emergence, selection and persistence of different lineages and genetic vectors of bla_KPC_. This information will improve our understanding of how to intervene to limit the transmission of drug-resistant Enterobacterales in healthcare settings, and avoid incorrect inference.

## 10. Author statements

### 10.3 Ethical approval

This study was performed as part of a Trust board-approved and UK Health Security Agency (formerly Public Health England) long-term outbreak response, and ethics approval was not required under NHS governance arrangements; all analyses were undertaken using anonymised data. This was reviewed and agreed by the Joint Research Office of the Oxford University Hospitals NHS Foundation Trust and the University of Oxford (who oversee research governance, ethics and assurance for our institution), and who deemed that neither sponsorship nor research ethics review was required.

## 10.4 Acknowledgements

We are grateful to the microbiology staff and infection prevention and control teams at Manchester University NHS Foundation Trust.

The Transmission of Carbapenemase-producing Enterobacteriaceae (TRACE) Investigators’ Group is made up of the following (listed alphabetically, includes several of the authors also listed by name in the main author list): Zoie Aiken, Oluwafemi Akinremi, Aiysha Ali, Julie Cawthorne, Paul Cleary, Derrick W. Crook, Valerie Decraene, Andrew Dodgson, Michel Doumith, Matthew J. Ellington, Ryan George, John Grimshaw, Malcolm Guiver, Robert Hill, Katie L. Hopkins, Rachel Jones, Cheryl Lenney, Amy J. Mathers, Ashley McEwan, Ginny Moore, Andrew Mumford, Mark Neilson, Sarah Neilson, Tim E.A. Peto, Hang T.T. Phan, Mark Regan, Anna C. Seale, Nicole Stoesser, Jay Turner-Gardner, Vicky Watts, A. Sarah Walker, Jimmy Walker, David H. Wyllie, William Welfare and Neil Woodford.

