## Supplementary material for "Genomic epidemiology and longitudinal sampling of ward wastewater environments and patients reveals complexity of the transmission dynamics of bla_KPC_-carbapenemase-producing Enterobacterales in a hospital setting"

Supplementary methods

*Environmental sampling - details*

Wastewater site types sampled sampled were:

- Clinical handwash basin drains (sites used specifically for staff handwashing in patient bays)
- Handwash basin drains (handwash basins for patient use in en-suite bathrooms or ward toilets)
- Mop sinks (into which discarded water following the use of mops for cleaning was disposed of)
- Sluices/sluice sinks (into which patient bodily fluids/waste was disposed of)
- Utility sinks (in utility settings, but not used specifically for waste disposal)
- Toilets
- Shower/bath drains.

Sink P-traps, shower drains or toilet water aspirates were centrifuged at 4000rpm for 10mins, 15mls of supernatant were discarded, and the pellet re-suspended in the remaining 5mls. One ml of sample was incubated in 5mls trypticase soy broth with an ertapenem disc aerobically at 37°C overnight.

*Patient rectal screen processing - details*

The Cepheid Xpert Carba-R assay was used for rapid testing on specimens from patients with admissions to the Trust in the past 12 months, those admitted from overseas, or transfers to other hospitals; the in-house multiplex assay was used for other sample types to mitigate cost of testing.

*Species-specific references used for sequence read mapping*

*Citrobacter freundii* (GenBank accession: CP011612.1) - 4,976,908bp

*Enterobacter cloacae* (CP001918.1) - 5,314,581bp

*Escherichia coli* (AE014075.1) - 5,231,428bp

*Klebsiella pneumoniae* (CP000647.1) - 5,315,120bp

*Raoultella ornithinolytica* (NC_021066.1) - 5,398,151bp

*Variant calling, filtering, generation of consensus fasta sequences*

Prior to reference-based mapping Illumina data were trimmed using cutadapt (version 1.5). Repetitive regions of the reference were identified using self-self BLASTn analysis with default settings (ncbi-blast-2.2.23+); these regions were then masked prior to mapping and base calling. Properly paired sequence reads were mapped to the reference using Stampy (v1.0.23)

Single-nucleotide variants (SNVs) were determined across all mapped non-repetitive sites using SAMtools (vsamtools-1.4.1) mpileup. mpileup was run twice to separate high-quality base calls from low-quality base calls; variant call format (VCF) files of annotated variant sites were created using vcftools (v0.1.9). Base calls derived from VCF files were filtered to retain only high quality calls using the following criteria:

- The proportion of high-quality bases supporting the call was ≥90%, and ≥5
  high-quality bases were required as a minimum, with at least one in the
  forward direction and at least one in the reverse direction
- The root of the mean square mapping quality of reads covering the site was
  ≥30
- The Phred scaled quality supporting the call was ≥25
- Reads spanning the site were made up of ≥35% high-quality bases
- The site was not called as heterozygous

Core variable sites (site called in all sequenced isolates, excluding “N” or “-” calls) derived from mapping to the reference were “padded” with invariant ACGT nucleotides in a proportion consistent with the ACGT content and length of the reference genome to generate a modified alignment of input sequences to generate phylogenies.

*Detail of the Gower’s distance estimation for isolates based on species-strain, plasmid replicon, IS/AMR gene profiles, Tn4401 sequence and Tn4401 target site sequences*

For each of the genetic features considered and listed in the text, namely:

- the species-strain type (i.e. species cluster number defined on the basis of genomic relatedness evaluated in SNVs compared to a reference)
- the plasmid replicon profile - i.e. the combination of plasmid replicons identified for each sequence using PlasmidFinder,
- the insertion sequence (IS) profile - i.e. the combination of IS types identified in each sequence using ISFinder,
- the antimicrobial resistance gene profile - i.e. the combination of AMR genes identified in each sequence by comparing against the CARD database,
- the Tn*4401* and target site sequence (TSS) profiles, representing the 5bp R and L flank signatures around any Tn*4401* element identified, using TETyper,

we generated a composite profile combining all of these profiles. We then compared all these composite profiles for each pair of isolates using a Gower’s distance, which is an established dissimilarity coefficient that defines how different two records are. This distance is represented as a number between 0 (identical) and 1 (maximally dissimilar). The Gower distance calculation was implemented in the daisy packge in R, as described - further details are available here: <https://www.rdocumentation.org/packages/cluster/versions/2.1.2/topics/daisy>.

*Details of SCOTTI run for* KPC-K. *pneumoniae strain 9*

SCOTTI was run on an alignment of variable sites for isolates making up the KPC-*K. pneumoniae* 9 cluster, padded to the length of the reference genome. Isolates were assigned to hosts; the period of CPE colonisation “infectious period” was defined as the interval spanning any positive result bounded by negative screens. The maximum number of demes was represented by the number of people on the ward who were CPE-culture positive between the first KPC-*K. pneumoniae* 9 sample and the last. A sensitivity analysis using ward admission/discharge dates as the “infectious period” was also considered, and made little difference on the outputs.

*Details of alternative heuristic approach for inferring transmission based on accessory cluster assignation*

Following on from the SCOTTI analysis for KPC-*K. pneumoniae 9*, we clustered the pangenome by annotating assemblies for each isolate and then running the command panaroo -i */*.gff -o kpne9_panaroo --clean-mode strict. Accessory clusters were assigned as in Fig.8. For Fig.6, transmission links were inferred on the basis of the following approach:

- Temporal overlap on same ward most consistent with transmission
- Niche colonised with KPC- *K. pneumoniae 9* earliest most likely to be the originating source
- If negative screening culture later than a culture-positive case then most likely to be recipient

Appendix results

76 sites were sampled on the cardiac unit (1,564 sampling events; 08/Jan/2016-28/Dec/2016), 129 sites on the acute medicine unit (1,309 sampling events, 18/Jul/2016-27/Dec/2016), and 144 sites on the geratology unit (1615 sampling events, 18/Jul/2016-28/Dec/2016). Of these, 23/76 (30%) sites and 111/1,564 (7.1%) sampling events on the cardiac unit were positive for KPC-E, 38/129 (29%) sites and 86/1,309 (6.6%) sampling events on the acute medicine unit and 40/144 [28%] and 122/1,615 (7.6%) on the geratology unit.

5,913 patients were admitted to any of the study wards at least once during the study period (01/Jan/2016-31/Dec/2016), of which 5,601 (94.7%) had at least one specimen taken for microbiological culture. In total, 49,923 specimens were taken from these patients during the study time period.

**Table S1. Sink site location codes.**

| **Sample location code** | **Location descriptor** | **Ward** |
| --- | --- | --- |
| T1 | W3W4 F Staff WC Toilet | W3/W4 |
| T2 | W3W4 FStaff WC HWB Drain | W3/W4 |
| T3 | W3W4 M Staff WC Toilet | W3/W4 |
| T4 | W3W4 M Staff WC HWB Drain | W3/W4 |
| T5 | W3W4 Kitchen Utility Sink | W3 |
| T6 | W3W4 Kitchen HWB Drain | W3 |
| T7 | W3W4 Dom Rm CHWB Drain | W3/W4 |
| T8 | W3W4 Doms Rm Mop Sink upper | W3/W4 |
| T9 | W3 Entrance HWB Drain | W3 |
| T10 | W3 Day Rm CHWB Drain | W3 |
| T11 | W3 Side Rm CHWB Drain | W3 |
| T12 | W3 Side Rm CHWB Drain | W3 |
| T13 | W3 Side Rm CHWB Drain | W3 |
| T14 | W3 Side Rm Toilet | W3 |
| T15 | W3 Side Rm HWB Drain | W3 |
| T16 | W3 Bay CHWB Drain Left | W3 |
| T17 | W3 Bay CHWB Drain Right | W3 |
| T18 | W3 Bay CHWB Drain Left | W3 |
| T19 | W3 Bay CHWB Drain Right | W3 |
| T20 | W3 Side Rm CHWB Drain | W3 |
| T21 | W3 Side Rm Toilet | W3 |
| T22 | W3 Side Rm HWB Drain | W3 |
| T23 | W3 Bay CHWB Drain 1st | W3 |
| T24 | W3 Bay CHWB Drain 2nd | W3 |
| T25 | W3 Bay Toilet | W3 |
| T26 | W3 Bay HWB Drain | W3 |
| T27 | W3 Bay Shower Drain | W3 |
| T28 | W3 Shower Rm Shower Drain | W3 |
| T29 | W3 Shower Rm Toilet | W3 |
| T30 | W3 Shower Rm HWB Drain | W3 |
| T31 | W3 Dirty Util CHWB Drain | W3 |
| T32 | W3 Dirty Util Utility Sink | W3 |
| T33 | W3 Dirty Util Sluice | W3 |
| T34 | W3 Dirty Util Sluice Sink | W3 |
| T35 | W3 Ward WC Toilet | W3 |
| T36 | W3 Ward WC HWB Drain | W3 |
| T37 | W3 Shower Rm Shower Drain | W3 |
| T38 | W3 Shower Rm Toilet | W3 |
| T39 | W3 Shower Rm HWB Drain | W3 |
| T40 | W4 Entrance CHWB Drain | W4 |
| T41 | W3 Treatment Rm CHWB Drain | W3 |
| T42 | W3 Treatment Rm Utility Sink | W3 |
| T43 | W4 Treatment Rm CHWB Drain | W4 |
| T44 | W4 Treatment Rm Utility Sink | W4 |
| T45 | W4 Side Rm CHWB Drain | W4 |
| T46 | W4 Side Rm CHWB Drain | W4 |
| T47 | W4 Side Rm CHWB Drain | W4 |
| T48 | W4 Dirty Util CHWB Drain | W4 |
| T49 | W4 Side Rm HWB Drain | W4 |
| T50 | W4 Bay CHWB Drain Left | W4 |
| T51 | W4 Bay CHWB Drain Right | W4 |
| T52 | W4 Bay CHWB Drain Left | W4 |
| T53 | W4 Bay CHWB Drain Right | W4 |
| T54 | W4 Side Rm CHWB Drain | W4 |
| T55 | W4 Staff Rm CHWB Drain | W4 |
| T56 | W4 Side Rm HWB Drain | W4 |
| T57 | W4 Bay CHWB Drain 2nd | W4 |
| T58 | W4 Bay CHWB Drain 1st | W4 |
| T59 | W4 Bay Toilet | W4 |
| T60 | W4 Bay HWB Drain | W4 |
| T61 | W4 Bay Shower Drain | W4 |
| T62 | W4 Shower Rm Shower Drain | W4 |
| T63 | W4 Shower Rm Toilet | W4 |
| T64 | W4 Shower Rm HWB Drain | W4 |
| T65 | W4 Dirty Util Sluice | W4 |
| T66 | W4 Dirty Util Sluice Sink | W4 |
| T67 | W4 Side Rm Toilet | W4 |
| T68 | W4 Side Rm Toilet | W4 |
| T69 | W4 Ward WC Toilet | W4 |
| T70 | W4 Ward WC HWB Drain | W4 |
| T71 | W4 Shower Rm Shower Drain | W4 |
| T72 | W4 Shower Rm Toilet | W4 |
| T73 | W4 Shower Rm HWB Drain | W4 |
| T74 | W4 Staff Rm Utility Sink Drain | W4 |
| T75 | W4 Dirty Util Utility Sink | W4 |
| T77 | W3W4 Dom Rm Mop Sink lower | W3/W4 |
| F1 | 45 46 Kitchen HWB Drain | W45/W46 |
| F2 | 45 46 Kitchen 1st Utility Sink | W45/W46 |
| F3 | 45 46 Kitchen 2nd Utility Sink | W45/W46 |
| F4 | 45 46 Disabled WC HWB Drain | W45/W46 |
| F5 | 45 46 Disabled WC Toilet | W45/W46 |
| F6 | 45 46 MF WC HWB Drain | W45/W46 |
| F7 | 45 46 MF WC Toilet | W45/W46 |
| F8 | 45 46 Staff Room Utility Sink | W45/W46 |
| F9 | 45 46 Staff Room CHWB Drain | W45/W46 |
| F10 | 45 46 Seminar Room CHWB Drain | W45/W46 |
| F11 | W45 CHWB Drain 1 | W45 |
| F12 | W45 CHWB Drain 2 | W45 |
| F13 | W45 HWB Drain | W45 |
| F14 | W45 Toilet | W45 |
| F15 | W45 Shower Drain | W45 |
| F16 | W45 CHWB Drain | W45 |
| F17 | W45 HWB Drain | W45 |
| F18 | W45 Toilet | W45 |
| F19 | W45 Shower Drain | W45 |
| F20 | W45 CHWB Drain | W45 |
| F21 | W45 HWB Drain | W45 |
| F22 | W45 Toilet | W45 |
| F23 | W45 Shower Drain | W45 |
| F24 | W45 CHWB Drain | W45 |
| F25 | W45 HWB Drain | W45 |
| F26 | W45 Toilet | W45 |
| F27 | W45 Shower Drain | W45 |
| F28 | W45 CHWB Drain | W45 |
| F29 | W45 HWB Drain | W45 |
| F30 | W45 Toilet | W45 |
| F31 | W45 Shower Drain | W45 |
| F32 | W45 CHWB Drain | W45 |
| F33 | W45 HWB Drain | W45 |
| F34 | W45 Toilet | W45 |
| F35 | W45 Shower Drain | W45 |
| F36 | W45 CHWB Drain | W45 |
| F37 | W45 HWB Drain | W45 |
| F38 | W45 Toilet | W45 |
| F39 | W45 Shower Drain | W45 |
| F40 | W45 Bath Bath Drain | W45 |
| F41 | W45 Bath HWB Drain | W45 |
| F42 | W45 Bath Toilet | W45 |
| F43 | W45 Pantry HWB Drain | W45 |
| F44 | W45 Pantry Utility Sink Drain | W45 |
| F45 | W45 Drug Room CHWB Drain | W45 |
| F46 | W45 Drug Room Utility Sink | W45 |
| F47 | W45 Treatment Room CHWB Drain | W45 |
| F48 | W45 Dirty Utility CHWB Drain | W45 |
| F49 | W45 Dirty Utility Sluice | W45 |
| F50 | W45 Dirty Utility Sluice Sink | W45 |
| F51 | W45 Shower Room HWB Drain | W45 |
| F52 | W45 Shower Room Shower Drain | W45 |
| F53 | W45 Dom Room Mop Sink Upper | W45 |
| F54 | W45 Dom Room Mop Sink Lower | W45 |
| F55 | W45 CHWB Drain | W45 |
| F56 | W45 HWB Drain | W45 |
| F57 | W45 Toilet | W45 |
| F58 | W45 Shower Drain | W45 |
| F59 | W45 CHWB Drain | W45 |
| F60 | W45 HWB Drain | W45 |
| F61 | W45 Toilet | W45 |
| F62 | W45 Shower Drain | W45 |
| F63 | W45 CHWB Drain | W45 |
| F64 | W45 HWB Drain | W45 |
| F65 | W45 Toilet | W45 |
| F66 | W45 Shower Drain | W45 |
| F67 | W45 CHWB Drain | W45 |
| F68 | W45 HWB Drain | W45 |
| F69 | W45 Toilet | W45 |
| F70 | W45 Shower Drain | W45 |
| F71 | W45 CHWB Drain | W45 |
| F72 | W45 HWB Drain | W45 |
| F73 | W45 Toilet | W45 |
| F74 | W45 Shower Drain | W45 |
| F75 | W45 CHWB Drain | W45 |
| F76 | W45 HWB Drain | W45 |
| F77 | W45 Toilet | W45 |
| F78 | W45 Shower Drain | W45 |
| F79 | W46 CHWB Drain 1 | W46 |
| F80 | W46 CHWB Drain 2 | W46 |
| F81 | W46 HWB Drain | W46 |
| F82 | W46 Toilet | W46 |
| F83 | W46 Shower Drain | W46 |
| F84 | W46 CHWB Drain | W46 |
| F85 | W46 HWB Drain | W46 |
| F86 | W46 Toilet | W46 |
| F87 | W46 Shower Drain | W46 |
| F88 | W46 CHWB Drain | W46 |
| F89 | W46 HWB Drain | W46 |
| F90 | W46 Toilet | W46 |
| F91 | W46 Shower Drain | W46 |
| F92 | W46 CHWB Drain | W46 |
| F93 | W46 HWB Drain | W46 |
| F94 | W46 Toilet | W46 |
| F95 | W46 Shower Drain | W46 |
| F96 | W46 CHWB Drain | W46 |
| F97 | W46 HWB Drain | W46 |
| F98 | W46 Toilet | W46 |
| F99 | W46 Shower Drain | W46 |
| F100 | W46 CHWB Drain | W46 |
| F101 | W46 HWB Drain | W46 |
| F102 | W46 Toilet | W46 |
| F103 | W46 Shower Drain | W46 |
| F104 | W46 CHWB Drain | W46 |
| F105 | W46 HWB Drain | W46 |
| F106 | W46 Toilet | W46 |
| F107 | W46 Shower Drain | W46 |
| F108 | W46 Bath Bath Drain | W46 |
| F109 | W46 Bath HWB Drain | W46 |
| F110 | W46 Bath Toilet | W46 |
| F111 | W46 Pantry HWB Drain | W46 |
| F112 | W46 Pantry Utility Sink Drain | W46 |
| F113 | W46 Drug Room CHWB Drain | W46 |
| F114 | W46 Drug Room Utility Sink | W46 |
| F115 | W46 Treatment Room CHWB Drain | W46 |
| F116 | W46 Dirty Utility CHWB Drain | W46 |
| F117 | W46 Dirty Utility Sluice | W46 |
| F118 | W46 Dirty Utility Sluice Sink | W46 |
| F119 | W46 Shower Room HWB Drain | W46 |
| F120 | W46 Shower Room Shower Drain | W46 |
| F121 | W46 Dom Room Mop Sink upper | W46 |
| F122 | W46 Dom Room Mop Sink lower | W46 |
| F123 | W46 CHWB Drain | W46 |
| F124 | W46 HWB Drain | W46 |
| F125 | W46 Toilet | W46 |
| F126 | W46 Shower Drain | W46 |
| F127 | W46 CHWB Drain | W46 |
| F128 | W46 HWB Drain | W46 |
| F129 | W46 Toilet | W46 |
| F130 | W46 Shower Drain | W46 |
| F131 | W46 CHWB Drain | W46 |
| F132 | W46 HWB Drain | W46 |
| F133 | W46 Toilet | W46 |
| F134 | W46 Shower Drain | W46 |
| F135 | W46 CHWB Drain | W46 |
| F136 | W46 HWB Drain | W46 |
| F137 | W46 Toilet | W46 |
| F138 | W46 Shower Drain | W46 |
| F139 | W46 CHWB Drain | W46 |
| F140 | W46 HWB Drain | W46 |
| F141 | W46 Toilet | W46 |
| F142 | W46 Shower Drain | W46 |
| F143 | W46 CHWB Drain | W46 |
| F144 | W46 HWB Drain | W46 |
| F145 | W46 Toilet | W46 |
| F146 | W46 Shower Drain | W46 |
| A1 | AM1 Staff Room Utility Sink | AM1 |
| A2 | AM1 CHWB | AM1 |
| A3 | AM1 Toilet | AM1 |
| A4 | AM1 HWB Drain | AM1 |
| A5 | AM1 Shower Drain | AM1 |
| A6 | AM1 CHWB | AM1 |
| A7 | AM1 Toilet | AM1 |
| A8 | AM1 HWB Drain | AM1 |
| A9 | AM1 Shower Drain | AM1 |
| A10 | AM1 CHWB | AM1 |
| A11 | AM1 Toilet | AM1 |
| A12 | AM1 HWB Drain | AM1 |
| A13 | AM1 Shower Drain | AM1 |
| A14 | AM1 CHWB | AM1 |
| A15 | AM1 Toilet | AM1 |
| A16 | AM1 HWB Drain | AM1 |
| A17 | AM1 Shower Drain | AM1 |
| A18 | AM1 CHWB | AM1 |
| A19 | AM1 Toilet | AM1 |
| A20 | AM1 HWB Drain | AM1 |
| A21 | AM1 Shower Drain | AM1 |
| A22 | AM1 CHWB | AM1 |
| A23 | AM1 Toilet | AM1 |
| A24 | AM1 HWB Drain | AM1 |
| A25 | AM1 Shower Drain | AM1 |
| A26 | AM1 CHWB Entry | AM1 |
| A27 | AM1 CHWB Room | AM1 |
| A28 | AM1 Toilet | AM1 |
| A29 | AM1 HWB Drain | AM1 |
| A30 | AM1 Shower Drain | AM1 |
| A31 | AM2 CHWB Room | AM2 |
| A32 | AM1 Staff WC Toilet | AM1 |
| A33 | AM1 Staff WC HWB Drain | AM1 |
| A34 | AM1 MF WC Toilet | AM1 |
| A35 | AM1 MF WC HWB Drain | AM1 |
| A36 | AM1 Kitchen 1st Utility Sink | AM1 |
| A37 | AM1 Kitchen 2nd Utility Sink | AM1 |
| A38 | AM1 Kitchen HWB Drain | AM1 |
| A39 | AM1 CHWB | AM1 |
| A40 | AM1 Toilet | AM1 |
| A41 | AM1 HWB Drain | AM1 |
| A42 | AM1 Shower Drain | AM1 |
| A43 | AM1 CHWB | AM1 |
| A44 | AM1 Toilet | AM1 |
| A45 | AM1 HWB Drain | AM1 |
| A46 | AM1 Shower Drain | AM1 |
| A47 | AM1 CHWB | AM1 |
| A48 | AM1 Toilet | AM1 |
| A49 | AM1 HWB Drain | AM1 |
| A50 | AM1 Shower Drain | AM1 |
| A51 | AM1 CHWB | AM1 |
| A52 | AM1 Toilet | AM1 |
| A53 | AM1 HWB Drain | AM1 |
| A54 | AM1 Shower Drain | AM1 |
| A55 | AM1 CHWB | AM1 |
| A56 | AM1 Toilet | AM1 |
| A57 | AM1 HWB Drain | AM1 |
| A58 | AM1 Shower Drain | AM1 |
| A59 | AM1 Drug Room HWB | AM1 |
| A60 | AM1 Treatment Room CHWB | AM1 |
| A61 | AM1AM2 Dirty Util Sluice | AM1/AM2 |
| A62 | AM1AM2 Dirty Util Sluice Sink | AM1/AM2 |
| A63 | AM1AM2 Dirty Util CHWB Drain | AM1/AM2 |
| A64 | AM2 Visitor Toilet Toilet | AM2 |
| A65 | AM2 Visitor Toilet HWB Drain | AM2 |
| A66 | AM2 Staff Room Utility Sink | AM2 |
| A67 | AM2 CHWB | AM2 |
| A68 | AM2 Toilet | AM2 |
| A69 | AM2 HWB Drain | AM2 |
| A70 | AM2 Shower Drain | AM2 |
| A71 | AM2 CHWB | AM2 |
| A72 | AM2 Toilet | AM2 |
| A73 | AM2 HWB Drain | AM2 |
| A74 | AM2 Shower Drain | AM2 |
| A75 | AM2 CHWB | AM2 |
| A76 | AM2 Toilet | AM2 |
| A77 | AM2 HWB Drain | AM2 |
| A78 | AM2 Shower Drain | AM2 |
| A79 | AM2 CHWB | AM2 |
| A80 | AM2 Toilet | AM2 |
| A81 | AM2 HWB Drain | AM2 |
| A82 | AM2 Shower Drain | AM2 |
| A83 | AM2 CHWB | AM2 |
| A84 | AM2 Toilet | AM2 |
| A85 | AM2 HWB Drain | AM2 |
| A86 | AM2 Shower Drain | AM2 |
| A87 | AM2 CHWB | AM2 |
| A88 | AM2 Toilet | AM2 |
| A89 | AM2 HWB Drain | AM2 |
| A90 | AM2 Shower Drain | AM2 |
| A91 | AM2 CHWB Entry | AM2 |
| A92 | AM2 CHWB Room | AM2 |
| A93 | AM2 Toilet | AM2 |
| A94 | AM2 HWB Drain | AM2 |
| A95 | AM2 Shower Drain | AM2 |
| A96 | AM2 CHWB Entry | AM2 |
| A97 | AM2 CHWB Room | AM2 |
| A98 | AM2 Toilet | AM2 |
| A99 | AM2 HWB Drain | AM2 |
| A100 | AM2 Shower Drain | AM2 |
| A101 | AM2 Staff WC Toilet | AM2 |
| A102 | AM2 Staff WC HWB Drain | AM2 |
| A103 | AM2 MF WC Toilet | AM2 |
| A104 | AM2 MF WC HWB Drain | AM2 |
| A105 | AM2 Kitchen 1st Utility Sink | AM2 |
| A106 | AM2 Kitchen 2nd Utility Sink | AM2 |
| A107 | AM2 Kitchen HWB Drain | AM2 |
| A108 | AM2 CHWB | AM2 |
| A109 | AM2 Toilet | AM2 |
| A110 | AM2 HWB Drain | AM2 |
| A111 | AM2 Shower Drain | AM2 |
| A112 | AM2 CHWB | AM2 |
| A113 | AM2 Toilet | AM2 |
| A114 | AM2 HWB Drain | AM2 |
| A115 | AM2 Shower Drain | AM2 |
| A116 | AM2 CHWB | AM2 |
| A117 | AM2 Toilet | AM2 |
| A118 | AM2 HWB Drain | AM2 |
| A119 | AM2 Shower Drain | AM2 |
| A120 | AM2 CHWB | AM2 |
| A121 | AM2 Toilet | AM2 |
| A122 | AM2 HWB Drain | AM2 |
| A123 | AM2 Shower Drain | AM2 |
| A124 | AM2 CHWB | AM2 |
| A125 | AM2 Toilet | AM2 |
| A126 | AM2 HWB Drain | AM2 |
| A127 | AM2 Shower Drain | AM2 |
| A128 | AM2 Drug Room HWB | AM2 |
| A129 | AM2 Treatment Room CHWB | AM2 |

**Figure S2. Distribution of pairwise SNV (single nucleotide variant) distances between isolates amongst major CPE species identified.** SNVs are calculated from recombination-adjusted phylogenies. Plots demonstrate that our definition of strain was conservative, and that it some cases this could be further divided into sub-groupings based on SNV distributions; however, no single threshold was widely applicable.

**A. *Citrobacter freundii***

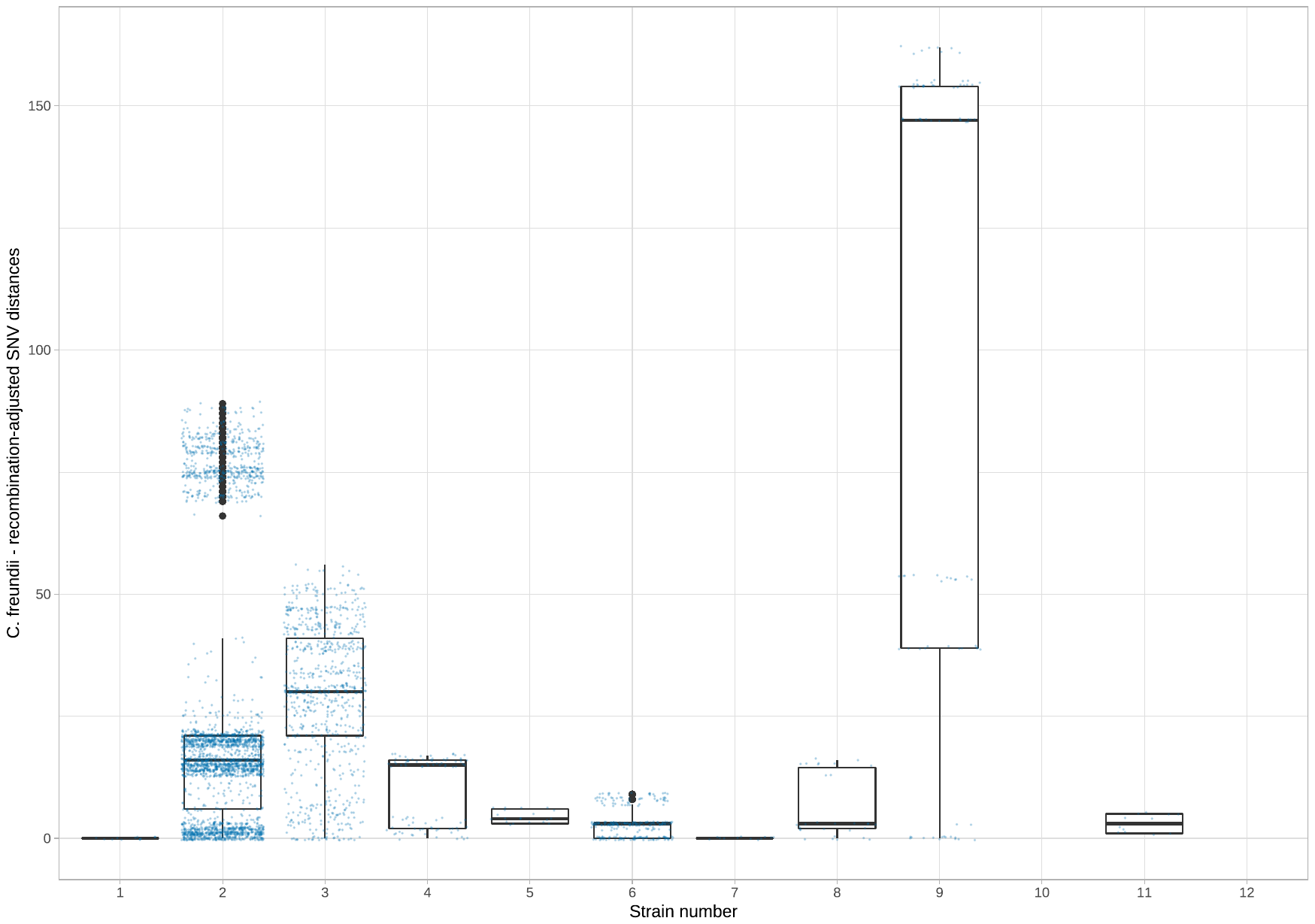

**B. *Enterobacter cloacae***

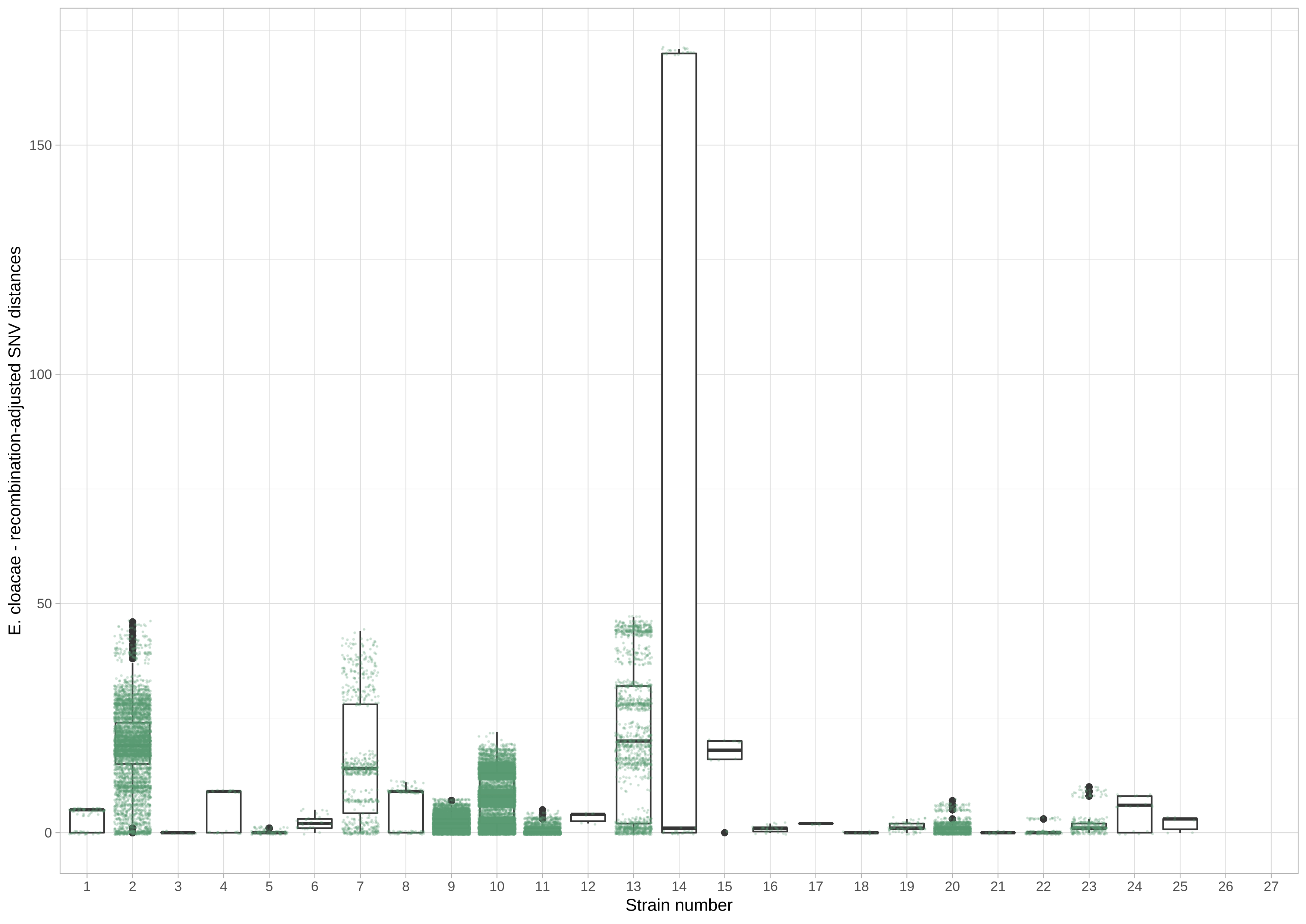

**C. *Escherichia coli***

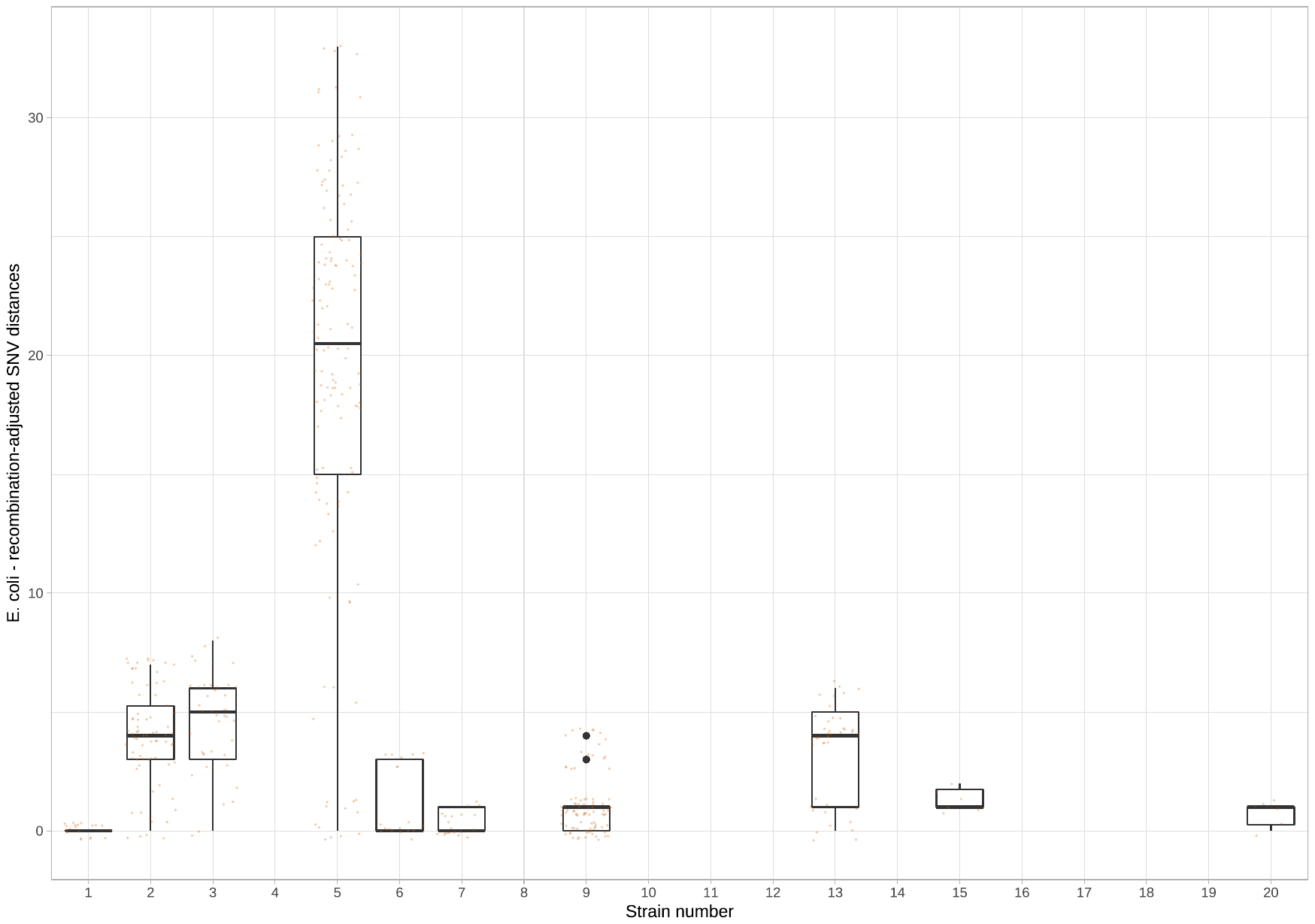

**D. *Klebsiella pneumoniae***

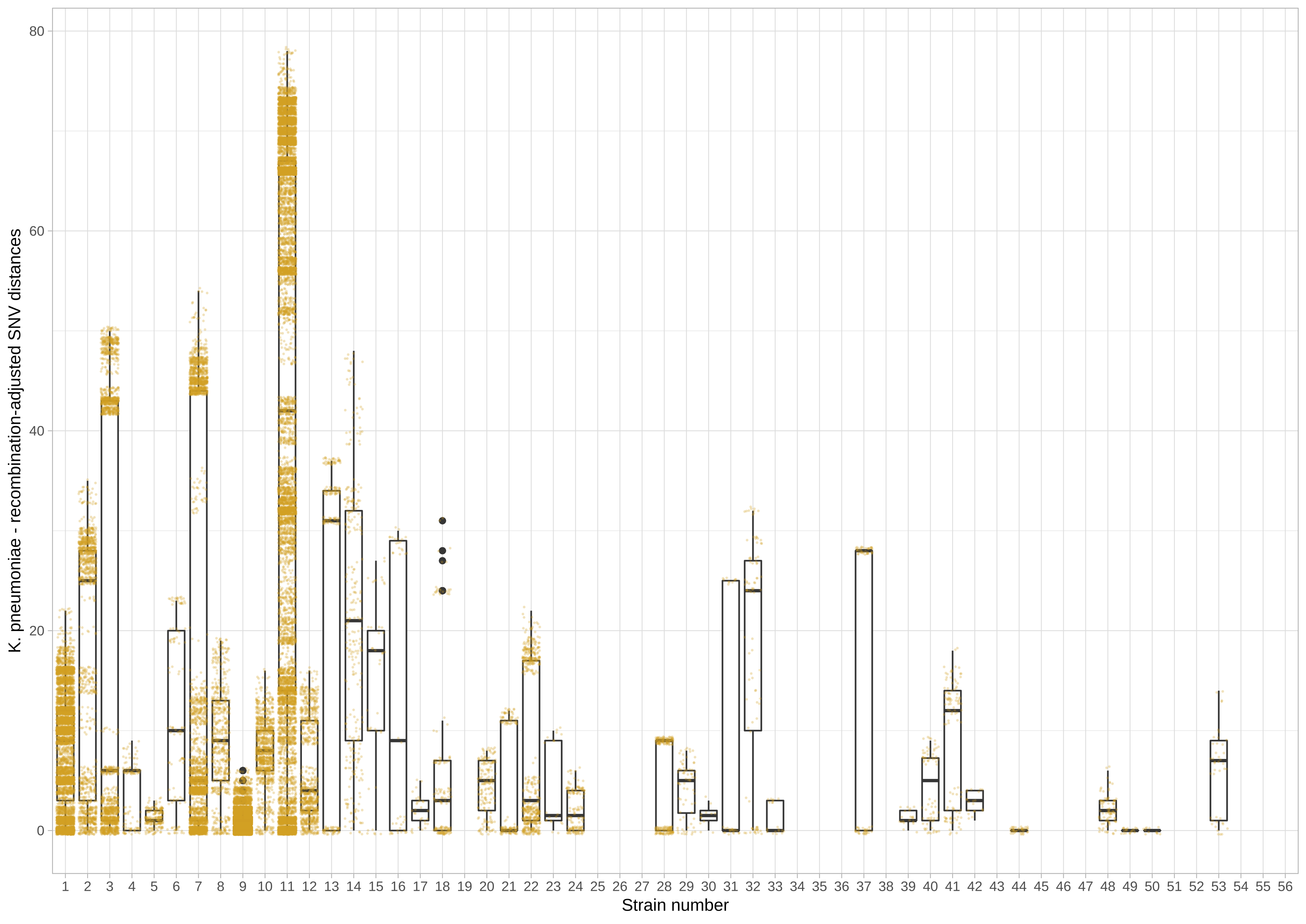

**E. *Raoultella ornithinolytica***

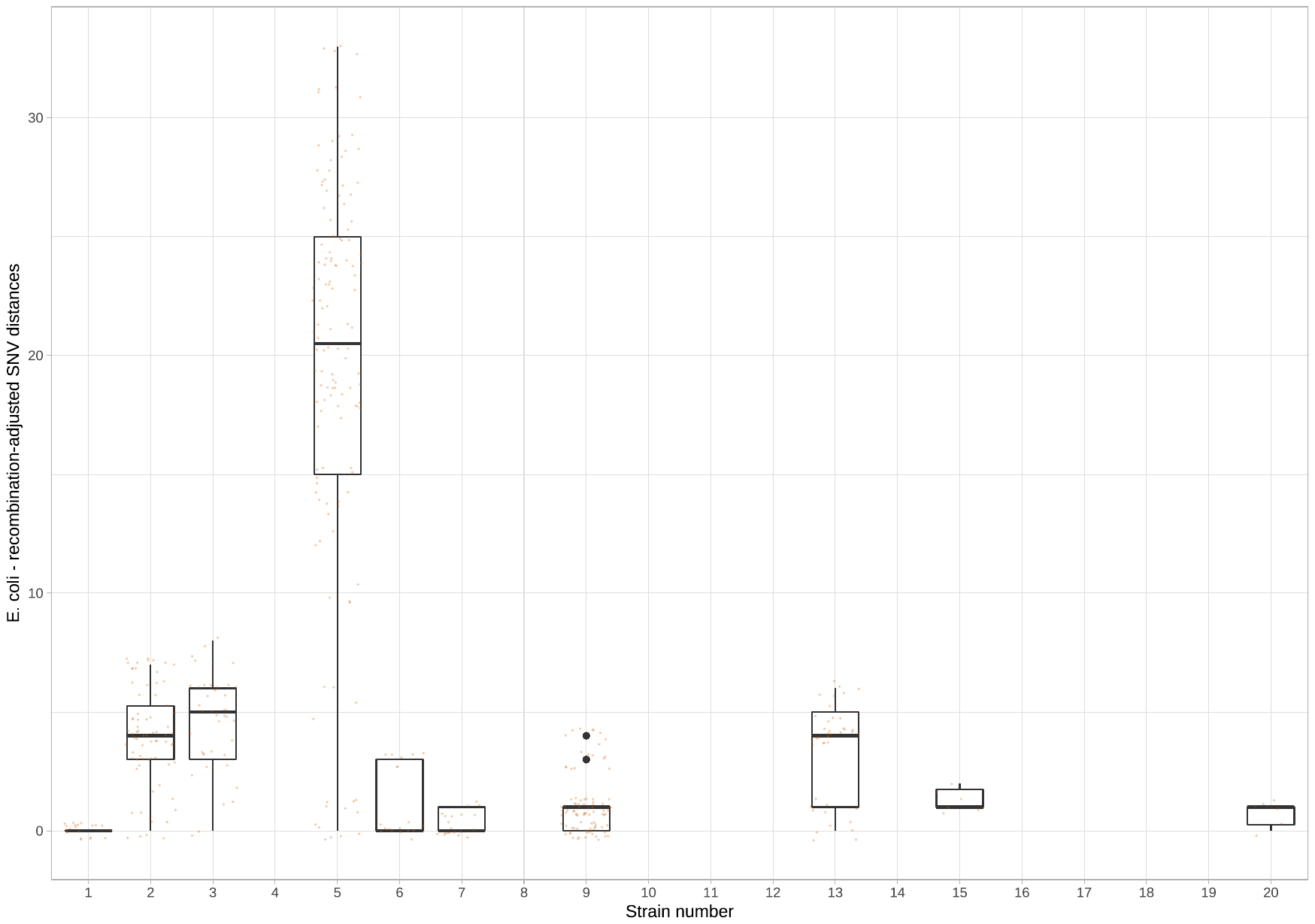

**Figure S3. Counts of KPC-E-positive and negative sampling events by environmental site, stratified by unit.** Note the different y-axis scales indicating the number of sampling events, related to the fact that the cardiology unit was sampled over a 12-month period and the other two units over a 6-month period. Red/top panel - acute medicine unit sites, blue/middle panel - geratology sites, green/bottom panel - cardiology unit sites.

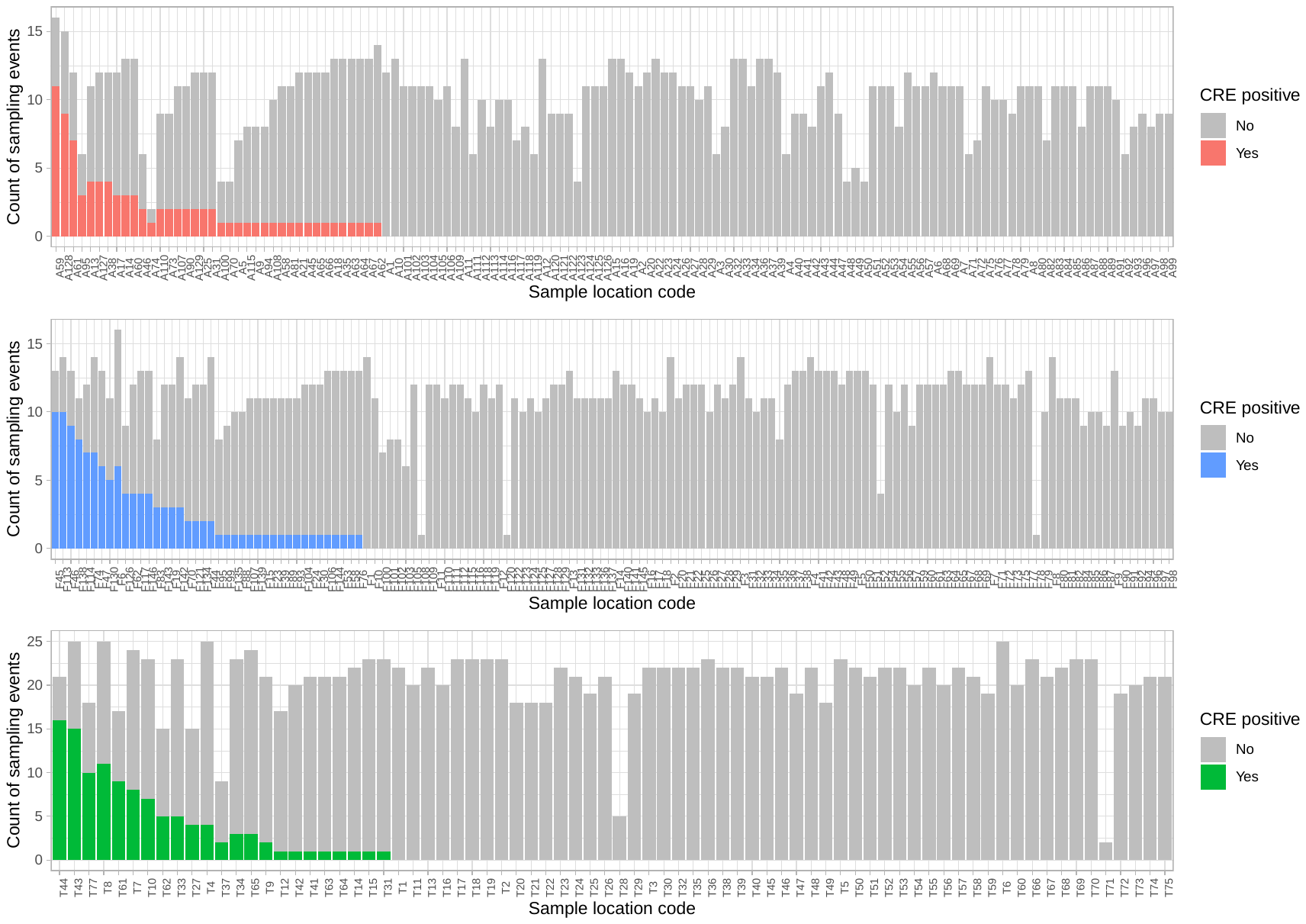

**Figure S4. Enterobacterales species cultured from all specimens taken from the study population during the study period, by carbapenem susceptibility and with species identification performed using routine laboratory methods.** The top panel shows all top ranked species; the bottom panel is an enlargement (altered y-scale) to enable better visualisation of species for which there are ≤150 isolates. *Enterobacter cloacae* complex includes: *E. cloacae* subsp *cloacae*, *E. asburiae*, *E. kobei*, *E. ludwigii*, *E. hormaechei*, *E. nimipressuralis*. Carbapenem susceptibility of 1=non-susceptible, 0=susceptible.

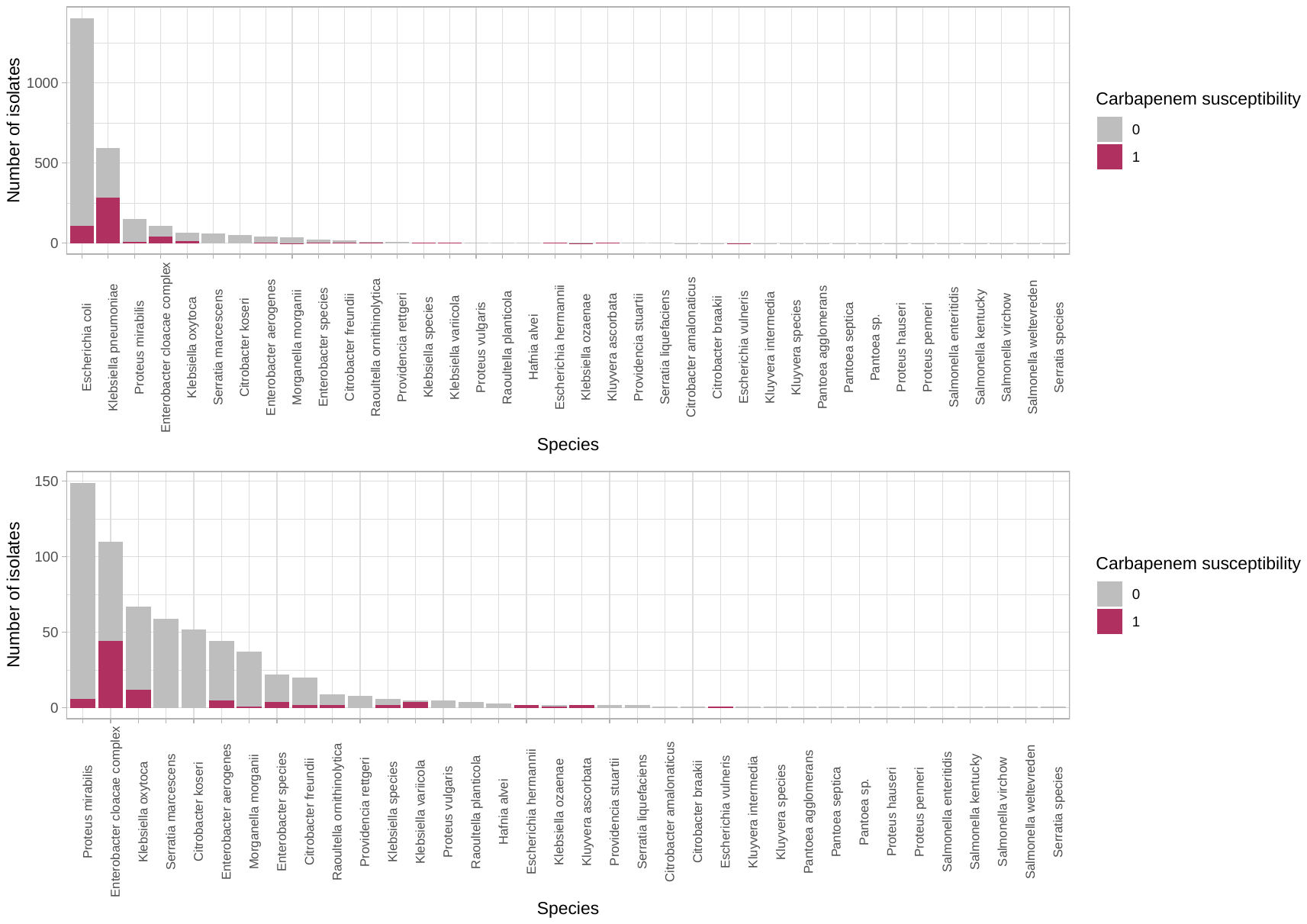

**Table S2. Distribution of major carbapenemases (targeted molecular testing performed for *bla*_NDM_, *bla*_KPC_ and *bla*_OXA-48_).** Details given for the subset of patient samples on which these data were available (n=344).

| **Single carbapenemase gene identified** | |
| --- | --- |
| **Carbapenemase gene** | **n** |
| *bla*_KPC_ | 296 |
| *bla*_OXA-48-like_ | 20 |
| *bla*_NDM_ | 11 |
| **Multiple carbapenemase genes identified** | |
| *bla*_NDM_ + *bla*_OXA-48-like_ | 10 |
| *bla*_KPC_ + *bla*_NDM_ | 1 |
| *bla*_KPC_ + *bla*_OXA-48-like_ | 5 |
| *bla*_KPC_ + *bla*_NDM_ + *bla*_OXA-48-like_ | 1 |

**Table S3. Sample type of all specimens cultured from the study population during the study period and Enterobacterales-positive status.** Data shown for 1^st^ January 2016-31^st^ December 2016 inclusive.

| **Sample type** | **Total samples** | **Negative/non-Enterobacterales culture result**  **n (% of samples by type)** | **Carbapenem-susceptible Enterobacterales isolates**  **n (% of Carbapenem-susceptible Enterobacterales isolates)** | **Carbapenem-resistant Enterobacterales isolates n (% Carbapenem-resistant Enterobacterales isolates)** |
| --- | --- | --- | --- | --- |
| Rectal screen | 26,875 | 26,520 (98.7%) | (12) (1.0%) | 387 (81.0%) |
| Urine | 8,404 | 6,972 (82.9%) | 1,388 (64.7%) | 52 (10.9%) |
| Wound or intra-abdominal swab | 6,190 | 5,930 (95.8%) | 306 (14.3%) | 13 (2.7%) |
| Blood culture | 4,102 | 3,959 (96.5%) | 154 (7.2%) | 4 (0.8%) |
| Respiratory sample | 2,830 | 2,588 (91.4%) | 256 (11.9%) | 21 (4.3%) |
| Other | 1,522 | 1,501 (98.6%) | 28 (1.3%) | 1 (0.2%) |
| **Total** | **49,923** | **47,470 (95.1%)** | **2,144** | **478** |

**Figure S5. Schematic of sharing between unique isolate niches with identical strain-level, plasmid replicon, Tn*4401*/TSS type, IS and AMR gene profiles.** Each niche is labelled with a unique identifier (i.e. a number for patients, and a letter-number combination for environmental sites). Triangles represent patients, circles environmental sites. Black bars represent admission dates for patient cases. Shape colour denotes unit - i.e. red=acute medicine, blue=geratology, green=cardiac. SNP distances between consecutive sequenced isolate pairs are shown on the right.

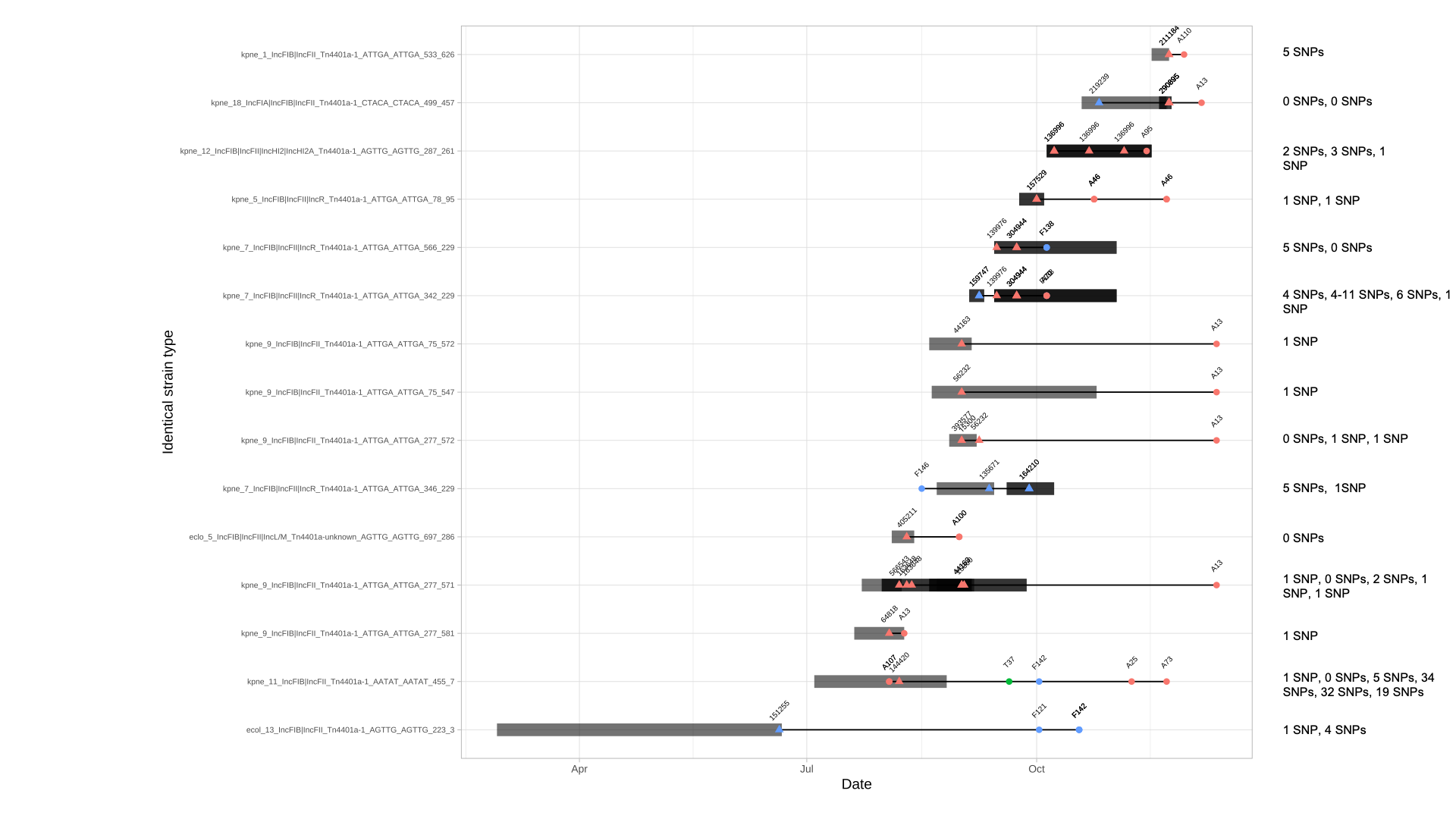

**Figure S6. Detailed breakdown of within-niche diversity.** Top panel: Within-patient KPC-E diversity. Pies reflect the proportion of strains within a sample that were identified using whole genome sequencing. “aher” = “*Escherichia hermanii*”, “cfre” = *Citrobacter freundii*, “eclo” = *Enterobacter cloacae*, “ecol” = *Escherichia coli*, “evul” = *Escherichia vulnificus*, “koxy” = *Klebsiella oxytoca*, “kpne”= *Klebsiella pneumoniae*, “lecl” = *Leclercia adecarboxylata*. Numbers after species abbreviations denote unique strains. Bottom panel: Within-wastewater sample KPC-E diversity. Pies reflect the proportion of strains within a sample that were identified using whole genome sequencing, with evaluation of diversity at the species-strain level. “aher” = *Escherichia hermanii*, “cfar” = *Citrobacter farmeri*, “easb” = *Enterobacter asburiae*, “pant” = *Pantoea* spp., “plur” = *Pluralibacter gergoviae*; other assignations as for top panel. Longitudinally positive samples are represented as separate pies along the y-axis.

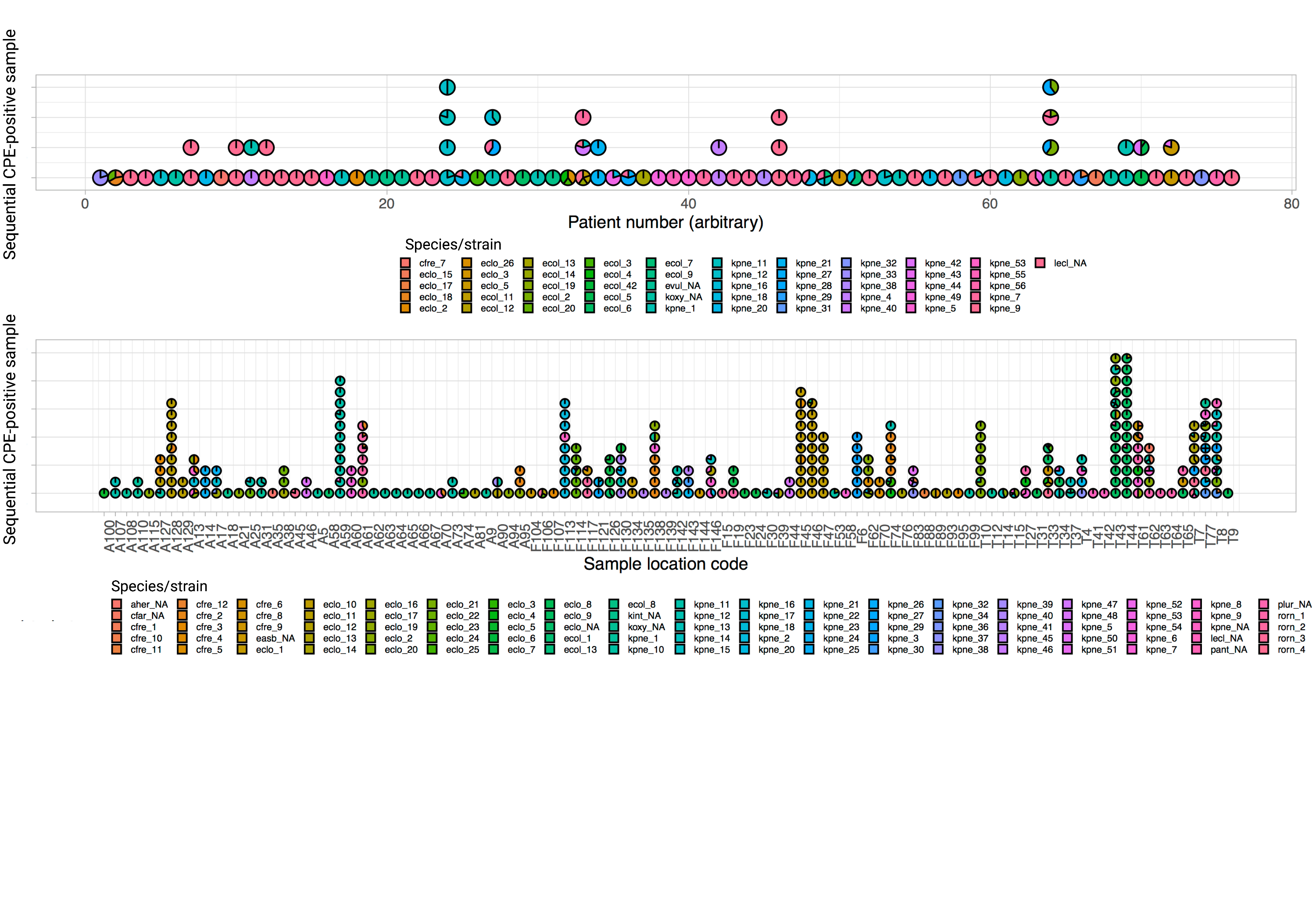

**Figure S7. *K. pneumoniae* strain 9 recombination-corrected phylogeny.**

Scale bar represents distance in SNVs. Labels from left to right represent: isolate name (*_<num>_<num> denotes sample, isolate and colony identifiers respectively), unit of sampling, date of sampling (year-month), niche, AMR gene profile, IS profile, plasmid finder profile, Tn*4401* type and 5bp target site sequences.

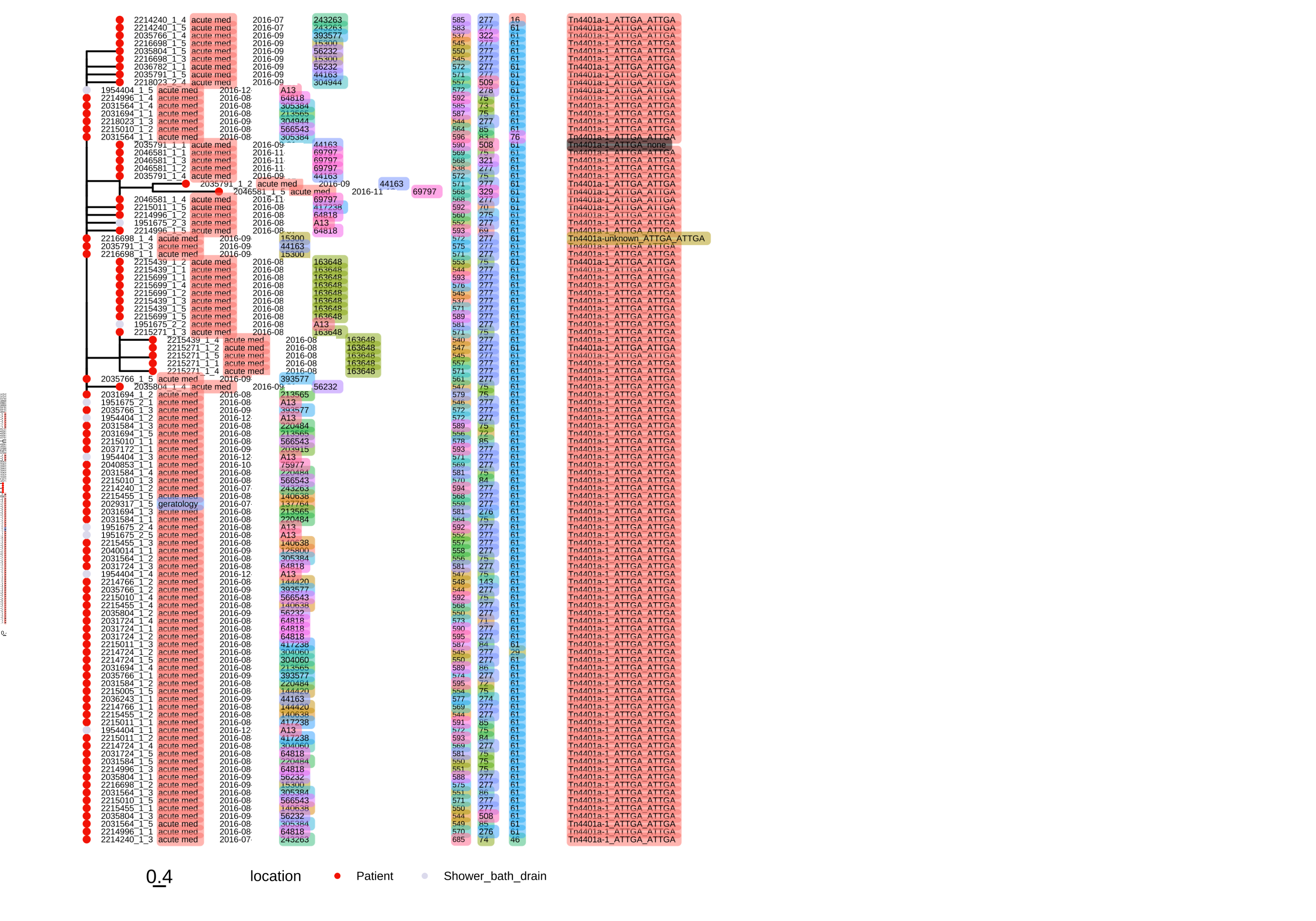

**Figure S8. *K. pneumoniae* strain 9 transmission network inferred by SCOTTI.** A) shows inferred direct transmission events, and B) inferred indirect transmission events. Each coloured blue shape and label represents a sample; arrows inferred transmission events and numbers alongside the arrows the posterior transmission probabilities.

A)

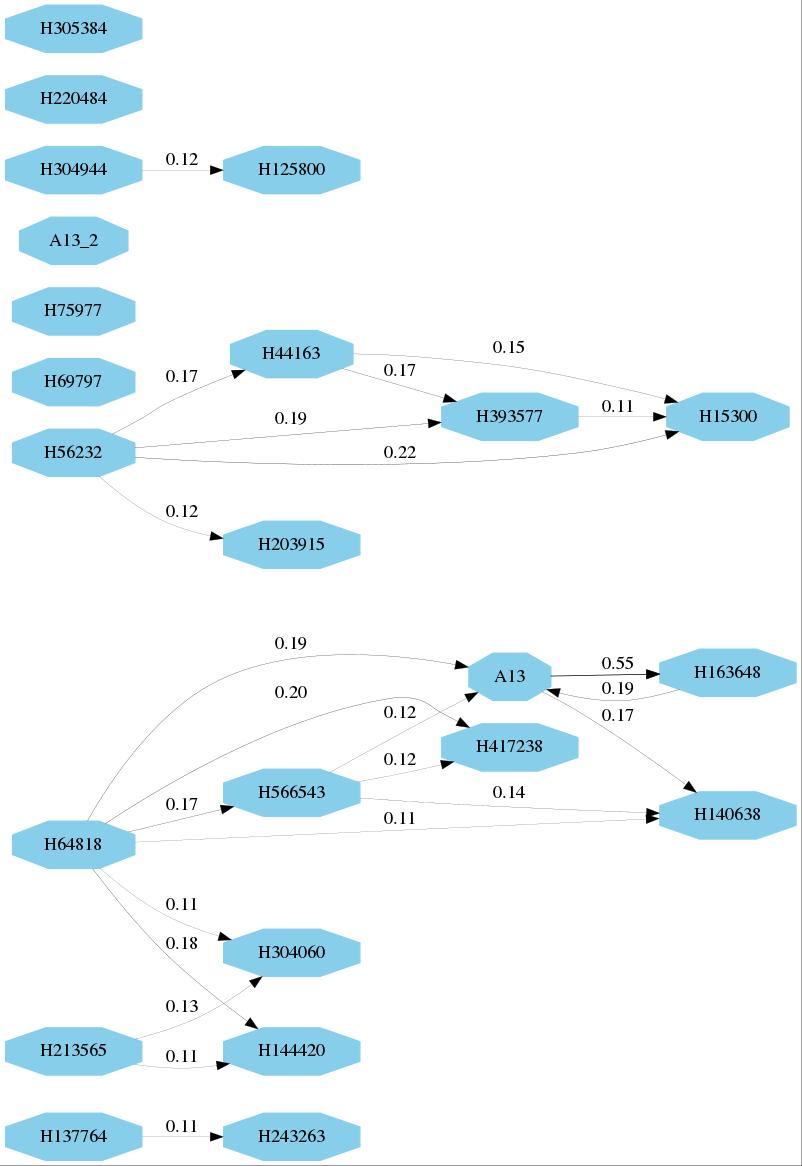

B)

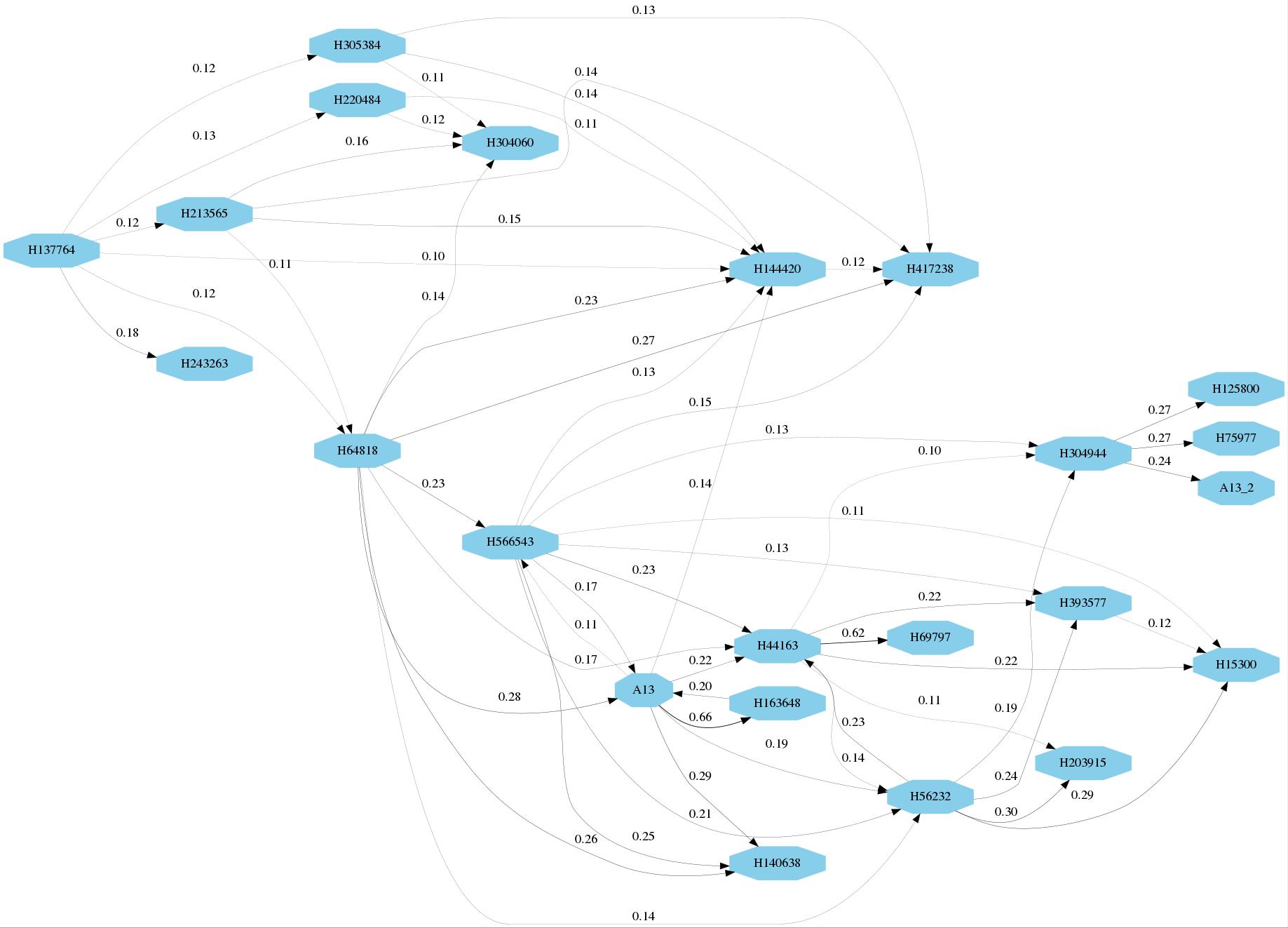

**Figure S9**. ***K. pneumoniae* strain 9 accessory gene presence/absence heatmap.** Derived using Panaroo. Each row represents an isolate; annotated CDSs in the non-core genome for the strain are shown along the x-axis. Black denotes presence, light grey absence of a particular CDS. Accessory genome CDS clusters are annotated on the dendrogram to the left of the heatmap.

**Figure S10. Two-way plot of the number of isolates for any given strain and the count of unique Tn*4401*-TSS types identified within that strain**. Higher counts of Tn*4401*-TSS types would be consistent with having either higher within-isolate transposition events or more frequent acquisition of other plasmids with different TSS contexts. Importantly, transposition events that result in the same TSS signature cannot be detected using this method.

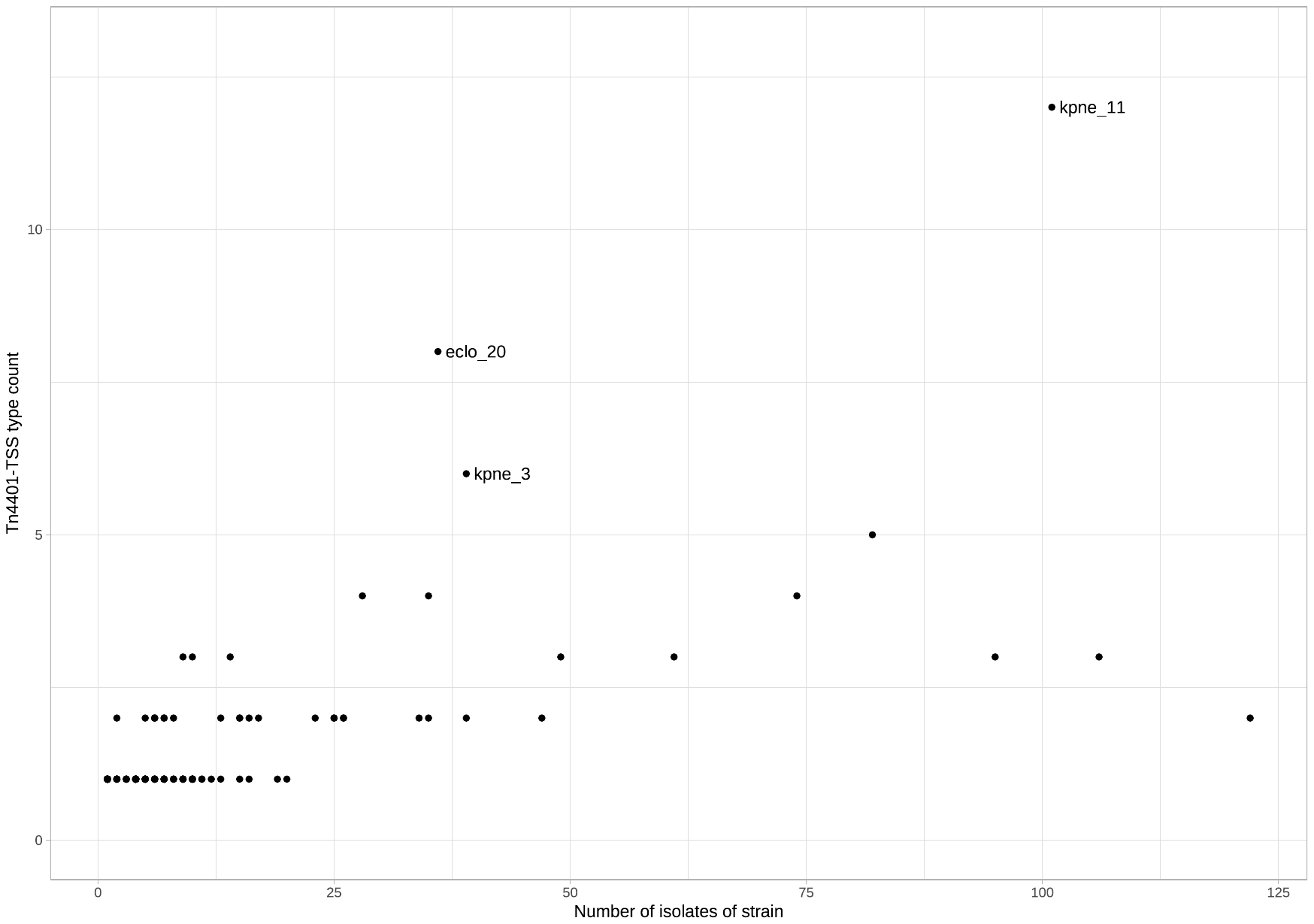

**Figure S11. *K. pneumoniae* strain 11 phylogeny demonstrating clusters separated by ~40SNVs**. Tn*4401* type and TSS flanking contexts observed are shown to the right of the phylogeny.

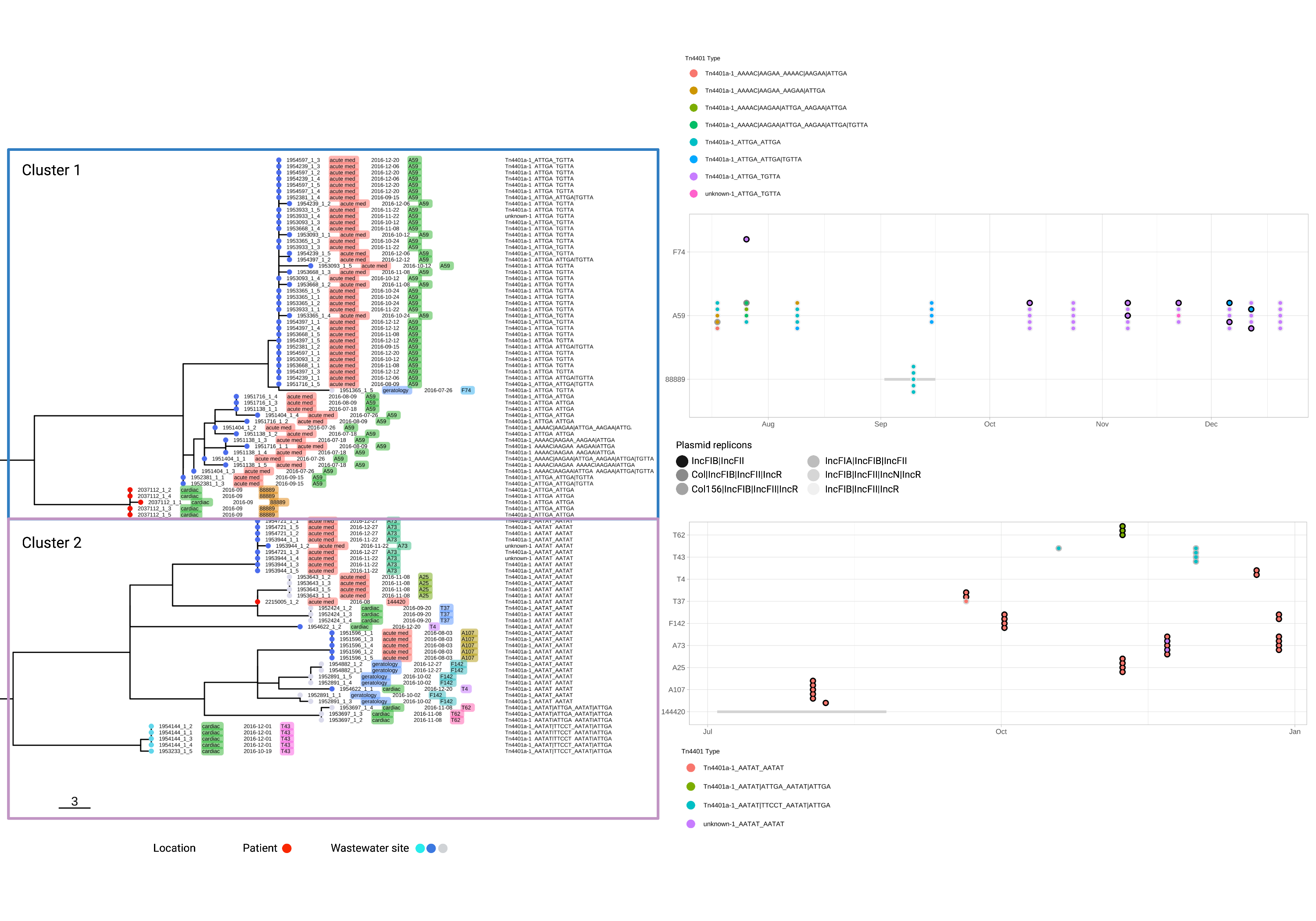
